# SARS-CoV-2 human challenge reveals single-gene blood transcriptional biomarkers that discriminate early and late phases of acute respiratory viral infections

**DOI:** 10.1101/2023.06.01.23290819

**Authors:** Joshua Rosenheim, Rishi K Gupta, Clare Thakker, Tiffeney Mann, Lucy CK Bell, Claire M Broderick, Kieran Madon, Loukas Papargyris, Pete Dayananda, Andrew J Kwok, James Greenan-Barrett, Helen R Wagstaffe, Emily Conibear, Joe Fenn, Seran Hakki, Rik GH Lindeboom, Lisa M Dratva, Briac Lemetais, Caroline M Weight, Cristina Venturini, Myrsini Kaforou, Michael Levin, Mariya Kalinova, Alex Mann, Andrew Catchpole, Julian C Knight, Marko Z. Nikolić, Sarah A. Teichmann, Ben Killingley, Wendy Barclay, Benjamin M Chain, Ajit Lalvani, Robert S Heyderman, Christopher Chiu, Mahdad Noursadeghi

## Abstract

Evaluation of host-response blood transcriptional signatures of viral infection have so far failed to test whether these biomarkers reflect different biological processes that may be leveraged for distinct translational applications. We addressed this question in the SARS-CoV-2 human challenge model. We found differential time profiles for interferon (IFN) stimulated blood transcriptional responses represented by measurement of single genes. MX1 transcripts correlated with a rapid and transient wave of type 1 IFN stimulated genes (ISG) across all cell types, which may precede PCR detection of replicative infection. Another ISG, IFI27, showed a delayed but sustained response restricted to myeloid peripheral blood mononuclear cells, attributable to gene and cell-specific epigenetic regulation. These findings were reproducible in diverse respiratory virus challenges, and in natural infection with SARS-CoV-2 or unselected respiratory viruses. The MX1 response achieved superior diagnostic accuracy in early infection, correlation with viral load and identification of virus culture positivity, with potential to stratify patients for time sensitive antiviral treatment. IFI27 achieved superior diagnostic accuracy across the time course of symptomatic infection. Compared to blood, measurement of these responses in nasal mucosal samples was less sensitive and did not discriminate between early and late phases of infection.

## Background

Host response biomarkers of viral infection have multiple potential clinical applications. These include diagnostic triage tests to direct prioritisation of confirmatory laboratory investigations, and to guide clinical management decisions with the aim of reducing unnecessary antibacterial prescribing, or directing infection control measures and antiviral treatment. Attention has mostly focussed on biomarker discovery in whole blood samples that enable easy and technically consistent access. Genome-wide transcriptional profiling has emerged as the most common unbiased data-driven approach due to the maturity of technical and analytical workflows^1^.

Numerous blood transcriptional signatures for host responses to viral infections have been identified in this way using case-control studies of natural infection or experimental viral challenge in humans, designed to discover the most parsimonious measurements that discriminate viral infections from healthy controls or other diseases. We previously tested the accuracy of such blood transcriptional signatures of viral infection, identified by systematic review, to detect incident SARS-CoV-2 infection^2^. We showed that the majority were highly correlated, and collectively driven by type 1 interferon (IFN) responses. Many, including single gene transcripts (such as that of IFI27) provided near perfect discrimination of PCR positive individuals compared to uninfected controls. In some, the transcriptional biomarkers identified infections before the first positive viral PCR in nasopharyngeal samples. The sensitivity of IFI27 measurements was further leveraged to provide evidence for abortive infections associated with virus specific T cell responses without detection of the virus by PCR^3^.

In observational studies of natural infection, it is not possible to synchronise the time course of exposure and replicative infection. This has precluded identification of temporally distinct host response biomarkers that may offer optimal solutions for different translational applications such as diagnostic triage or patient stratification for antiviral therapies. To address this limitation, we leveraged the first controlled human challenge model of SARS-CoV-2 infection, complemented with high frequency sampling to measure viral replication and host responses spanning the full time course of viral replication^4^. We updated our previous systematic review to undertake comprehensive head-to-head evaluation of all reported host transcriptional signatures of viral infection to date. We compared their ability to discriminate between groups of participants with and without evidence of replicative infection using whole blood samples stratified by time since experimental inoculation. For selected biomarkers, representative of differential host-responses over the time course, we evaluated associations with symptoms and viral load. We investigated their cellular source in single cell transcriptomic data, and the potential epigenetic mechanisms that may underpin their differential expression. We compared their measurement in blood and nasal swabs, and explored the extent to which our findings were generalisable to other respiratory viruses, in both experimental challenge and natural infection studies.

## Methods

### Research ethics

Regulatory approvals for the human studies presented herein were provided by the UK Health Research Authority under the following reference numbers: 20/UK/2001 and 20/UK/0002 for the SARS-CoV-2 challenge study; 20/NW/0231 for the INSTINCT study; 19/LO/1441 for the H3N2 influenza challenge study.

### Identification of blood transcriptional signatures of viral infection

We updated our previous systematic review of blood transcriptional biomarkers for viral infection^2^. In the current analysis, we amended our previous eligibility criteria to identify concise blood transcriptional signatures discovered or applied with a primary objective of diagnosis of viral infection from human whole-blood or peripheral blood mononuclear cell samples, excluding those exclusively intended to stratify severity of infection. Other eligibility criteria remained the same as our previous review. In our update, we searched MEDLINE for articles published up to 31 December 2022, using comprehensive MeSH and keyword terms for “viral infection”, “transcriptome”, “biomarker”, and “blood”, as previously^2^. Additional studies were identified in reference lists. Title and abstract screening was independently performed by two reviewers (CT and JGB); shortlisted articles were reviewed in full, with input from a third reviewer (RKG) to resolve conflicts. For eligible signatures, constituent genes, modelling approaches and gene weightings were extracted, with verification by a second reviewer. Multi-gene signatures are referred to using a prefix of the first-author’s name from the corresponding publication, and a suffix of the number of component genes. Single-gene signatures are referred to by the gene symbol.

### Human challenge and patient cohorts

The SARS-CoV-2 human challenge model in healthy seronegative volunteers has been described previously^4^. Briefly, 36 SARS-CoV-2 volunteers were inoculated intranasally with a standardized dose of D614G-containing pre-alpha wild-type SARS-CoV-2 under quarantine conditions. From 24 hours after inoculation, virus was quantified by PCR and culture in samples obtained at 12 hourly intervals from nose (mid-turbinate) and throat swabs for at least 14 days of quarantine, or longer if they remained in quarantine beyond 14 days because they still had detectable virus. A final sample was obtained at 28 days after challenge. Blood samples for RNA sequencing were collected into PAXgene tubes (Qiagen) before virus challenge, 6 hours after challenge, daily thereafter for 14 days and on day 28. Mid-turbinate MW013, X nose swabs (MW013, MedWire) for RNA sequencing were collected before virus challenge, and on days 1, 3, 5, 7, 10 and 14 after challenge, preserved in RNAprotect (Qiagen). Quantitation of viral load and symptoms has been described previously^4^. Two individuals who seroconverted in the interval between screening and inoculation were excluded from the present analysis, on the basis that they experienced a recent infection that may affect the biomarker expression that is the focus of this study.

The SARS-CoV-2 household contact study (INSTINCT) has been described previously^5^. Briefly, 52 household contacts of SARS-CoV-2 infected index cases recruited within 5 days of index case symptom onset provided nasopharyngeal swabs and blood RNA samples collected in PAXgene tubes on day of enrolment (day 0), day 7, day 14 and day 28. Nasopharyngeal swabs were used to measure viral copy number using PCR against the E-gene.

The Influenza H3N2 human challenge model has been described previously^6^. Briefly, 20 healthy volunteers were inoculated intranasally with a standardized dose of Influenza A/Belgium/4217/2015 (H3N2) under quarantine conditions. From 24 hours after inoculation, virus was quantified by PCR in nasal lavage samples obtained at 12 hourly intervals. Participants were ascertained to have replicative viral infection if found to have consecutive positive PCR tests at least 24 hours after challenge. Blood samples for RNA sequencing were collected into PAXgene tubes before virus challenge and days 1, 2, 3, 7, 10, 14, and 28 after challenge. Nasal curettage samples were collected on days −14 (baseline), 1, 2, 3, 7, 10 and 14 and preserved in TRIzol (ThermoFisher Scientific) as previously described^7^.

All RNA samples were stored at −80°C until processing.

### Transcriptional profiling

Total RNA was extracted from SARS-CoV-2 challenge PAXgene tubes using the PAXgene Blood RNA kit (Qiagen), including on-column DNase treatment and depleted of globin mRNA using the GLOBINclear Human Kit (Thermo Fisher Scientific). Total RNA was extracted from the INSTINCT SARS-CoV-2 household contact study and the H3N2 influenza challenge PAXgene tubes using the Qiasymphony PAXgene blood RNA kit, with subsequent DNase I treatment (Zymo) and clean-up using the RNA Clean and Concentrator-96 kit (Zymo), followed by globin mRNA and rRNA depletion using NEBNext® Globin & rRNA Depletion kits (New England BioLabs). Total RNA from SARS-CoV-2 challenge nasopharyngeal swabs and curettage samples was extracted using the RNeasy mini kit (Qiagen), including on-column DNase treatment. RNA concentrations were quantified using Qubit 2.0 Fluorometer (ThermoFisher Scientific). RNA integrity scores were determined using the Bioanalyser (RNA Nano 6000 Chip, Agilent) or 4200 Tape Station (Agilent).

Blood RNA samples from SARS-CoV-2 challenge underwent total RNA sequencing. DNA libraries were constructed using the KAPA RNA HyperPrep Kit with RiboErase (Roche) and sequenced on the Illumina NovaSemq 6000 platform using the NovaSeq 6000 S4 Reagent Kit (200 cycles) (Illumina), giving a median of 69.3 million (range 29.3-152.8) 100 base pair (bp) paired-end reads per sample. Nose swab RNA samples underwent mRNA sequencing. DNA libraries were constructed using the Kappa mRNA HyperPrep kit (Roche) and sequenced on the Illumina NextSeq platform the using the NextSeq 500/550 High Output Kit (75 cycles) (Illumina), giving a median of 32.4 million (range 3.2-176.2) 41bp paired-end reads per sample. Blood RNA samples from the INSTINCT SARS-CoV-2 household contact study underwent mRNA sequencing. DNA libraries were constructed using the NEBNext® Ultra™ II Directional RNA Library Prep Kit for Illumina (New England Biolabs) and sequenced on the Illumina HiSeq 4000 using the HiSeq 3000/4000 PE Cluster and SBS kits (Illumina), giving a median of 26.1 million (range 18.34-56.04) 75bp paired-end reads per sample. Nose curettage RNA samples from the H3N2 human challenge underwent mRNA sequencing. DNA libraries were constructed using the NEBNext® Ultra™ II Directional RNA Library Prep Kit for Illumina (New England Biolabs) and sequenced on the Illumina HiSeq 4000 using the HiSeq 3000/4000 SBS kit (Illumina), giving a median of 74.9 million (range 44.2-122) 75bp paired-end reads per sample. Whole blood RNA samples from the H3N2 influenza challenge underwent mRNA sequencing, DNA libraries were constructed with the NEBNext® Ultra II Directional RNA Library Prep Kit for Illumina (New England BioLabs) and sequenced on the Illumina NovaSeq 6000 platform using the NovaSeq 6000 S2 200 cycles Flowcell (Illumina), with a target of 40 million paired-end reads per sample.

SARS-CoV-2 challenge sequencing reads were mapped to the reference transcriptome (Ensembl Human GRCh38 release 108) using Kallisto (version 0.46.1)^8^. 371 blood RNA samples from 34 seronegative individuals gave a median of 28.7 million (range 12.2-73.5) mapped reads per sample. 99 nose swab RNA samples gave a median of 23.9 million (range 2.5-138.6) mapped reads per sample. Transcript-level output Deseq2 normalised counts and transcripts per million values were summed on gene level and annotated with Ensembl gene ID, gene name, and gene biotype using the tximport (version 1.20.0) and biomaRt (version 2.48.0) Bioconductor packages in R^9–13^.

Sequencing reads from the INSTINCT SARS-CoV-2 household contact study were mapped to the reference transcriptome (NCBI Human GRCh38.p13) using STAR aligner (version 2.7.1a)^14^. 134 blood RNA samples from 52 individuals gave a median of 14.37 million (range 7.30-37.67) mapped reads per sample. Read count matrices were generated using featureCounts from the Rsubread package^15^ and normalised using the variance stabilised transformation from the DESeq2 package.

For the sequencing reads of whole blood RNA from the H3N2 influenza challenge, quality control was performed using with FastQC (v 0.11.7; https://www.bioinformatics.babraham.ac.uk/projects/fastqc/) and adapter sequences were removed using Trimmomatic (v 0.36)^16^. The reads were mapped against hg38 reference genome using STAR aligner (v 2.7.1a). The featureCounts tool from Subread package (v 1.5.2) was used for transcript quantification. Computed gene counts were used for downstream analyses. Whole blood samples from 19 donors gave a median of 10.1 million (range 8.5-12.7) mapped reads per sample (read length = 100bp). Transcript-level output Deseq2 normalised counts were annotated with Ensembl gene ID and gene name using biomaRt (version 2.46.3) Bioconductor packages in R.Sequencing reads from the H3N2 challenge were mapped to the reference transcriptome (Ensembl Human GRCh38.p13) using STAR (version 2.7.10a). Nasal RNA samples from 17 donors gave a median of 21.4 million (range 3.32–34.7) mapped reads per sample. Transcript-level output Deseq2 normalised counts were annotated with Ensembl gene ID and gene name using biomaRt (version 2.52.0) Bioconductor packages in R.

For RNAseq datasets that were generated in more than one batch, processing batch effects were excluded by principal component analysis of all RNAseq data (Supplementary Figure 2). Additional genome-wide transcriptomic microarray data were derived from previously published experimental challenge datasets of other respiratory viruses (GEO accession: GSE73072)^17^ and from a natural infection study of respiratory viruses (GEO accession: GSE68310)^18^. In each case, we used log-2 transformed and normalised data matrices to quantify biomarker scores, standardized to baseline samples.

### Signature scores

Analyses were performed in R (version 4.0.2). Scores for candidate transcriptional signatures were calculated as per the original author’s descriptions using transcripts per million values, as previously. Where component genes could not be identified in the RNA sequencing dataset (for example due to genes being withdrawn from the reference transcriptome), these genes were excluded from calculations. Scores were standardised to Z scores by subtracting the mean and dividing by the standard deviation of pre-inoculation samples, and were multiplied by −1 for scores intended to decrease in the presence of viral infection. Discrimination of each signature for the outcome of replicative infection was calculated as the area under the receiver operating characteristic curve (AUROC), with 95% confidence intervals, and stratified by day since inoculation, using the pROC package in R^19^. Correlation between signatures and with viral loads was quantified as Spearman rank correlation coefficients.

### Analysis of ATACseq data

Publicly available ATAC (Assay for Transposase Accessible Chromatin) sequencing fastq datasets derived from unstimulated human monocytes, B-cells and CD4 T-effector cells (GEO accession: GSE118189, European Nucleotide Archive accession: PRJNA484801)^20^ were analysed with the nf-core ATAC-seq analysis pipeline (v2.0) curated in Nextflow^21, 22^, using default parameters. Adaptors were trimmed using trimgalore (v0.6.7) and reads were aligned to the reference genome (NCBI GRCh38) using BWA (v 0.7.17)^23^. Duplicate reads were identified using picard (v2.27.4)^24^. Reads were filtered using SAMtools (v1.16.1)^25^. BEDtools (v.2.30.0)^26^ was used to remove duplicates, reads mapping to blacklisted regions and mitochondrial DNA, multimappers, unmapped reads or those not marked as primary alignments. Replicate datasets were merged using picard for some downstream analyses. Normalised scaled bigWig files were created using BEDtools and tracks were visualised using Integrative Genomics Viewer (v2.16.0)^27^. Peak calling was performed using MACS2 (v2.2.7.1)^28^ in broadpeak mode. Peaks were annotated to gene features using HOMER (v4.11)^29^ and a consensus peak-set was generated using BEDtools. Matrices of reads falling within consensus peaks were generated using featureCounts from the subread package (v2.0.1)^15^ for quantitation.

Publicly available single-cell ATACseq data from the COMBAT consortium^30^ (EGAD00001007963; Zenodo: https://doi.org/10.5281/zenodo.6120249) were reanalysed for read counts per cell type in established COVID infection from hospitalised COVID patients. Data were processed as described in the original publication using the ArchR software package (v0.9.3)^31^. The sequencing reads at the *IFI27* locus were plotted per cell type with the plotBrowserTrack function.

## Results

### Blood transcriptional signatures of viral infection

We updated our previous systematic review of the literature, to identify 26 blood transcriptional signatures associated with viral infection (Supplementary Figure 1A, Supplementary Table 1)^32–56^. These included six single gene biomarkers. The remaining multigene signatures were made up of 2-47 constituent genes. The composition of these signatures was generally distinct, reflected by low Jaccard indices in a matrix of pairwise comparisons (Supplementary Figure 1B).

### Viral infection outcomes in the SARS-CoV-2 controlled human challenge model

34 SARS-CoV-2 seronegative healthy volunteers subjected to nasal inoculation of a standardized dose of SARS-CoV-2 divided into two groups with (N=18) and without (N=16) evidence of sustained replicative infection from 2 days after challenge (Figure 1). Although the individual viral load profiles were different in nose and throat swabs, both measurements segregated the same participants into two groups with and without replicative infection.

**Figure 1.**
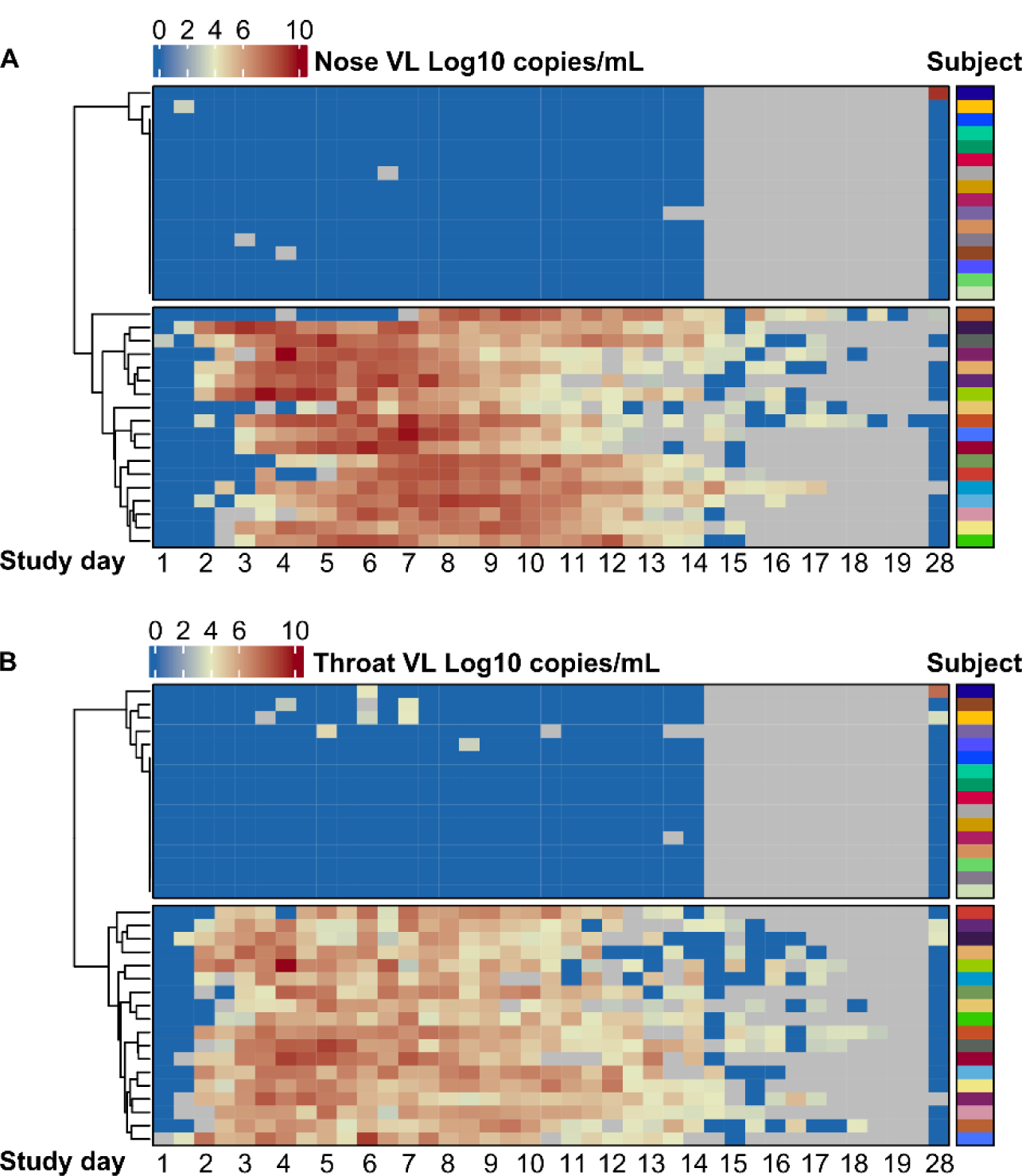
SARS-CoV-2 PCR viral load in nose and throat swabs following virus challenge. Quantitative viral load measurements by PCR from **(A)** nose and **(B)** throat swabs per participant (rows) stratified by time point (columns) after virus challenge, and clustered into two groups of participants with (N=18) and without (N=16) evidence of replicative virus replication. Grey colour denotes unavailable data points.

### Blood transcriptional biomarker discrimination of participants with and without sustained replicative SARS-CoV-2 infection

Blood transcriptional biomarker scores were calculated for each of the 26 signatures identified by systematic review, from RNA sequencing of whole blood samples at selected time points before and after viral inoculation (Figure 2A). Across this time course, all the biomarkers showed a transient increase in expression (Supplementary Figure 3) associated with replicative SARS-CoV-2 infection. We first ranked all biomarkers by their ability to discriminate between participants with and without replicative infection by area under the receiver operating characteristic curve (AUROC) across 14 days. We limited calculations to data from days 3, 7, 10 and 14, in order to achieve equal sampling frequency distribution across the time course of infection (Supplementary Figure 3). Point estimates of the AUROCs ranged between 0.6-0.99. 22 of the 26 biomarkers with point estimates ranging 0.92-0.99 were statistically comparable with overlapping 95% confidence intervals, suggesting most biomarkers were able to accurately discriminate participants with and without replicative infection.

**Figure 2.**
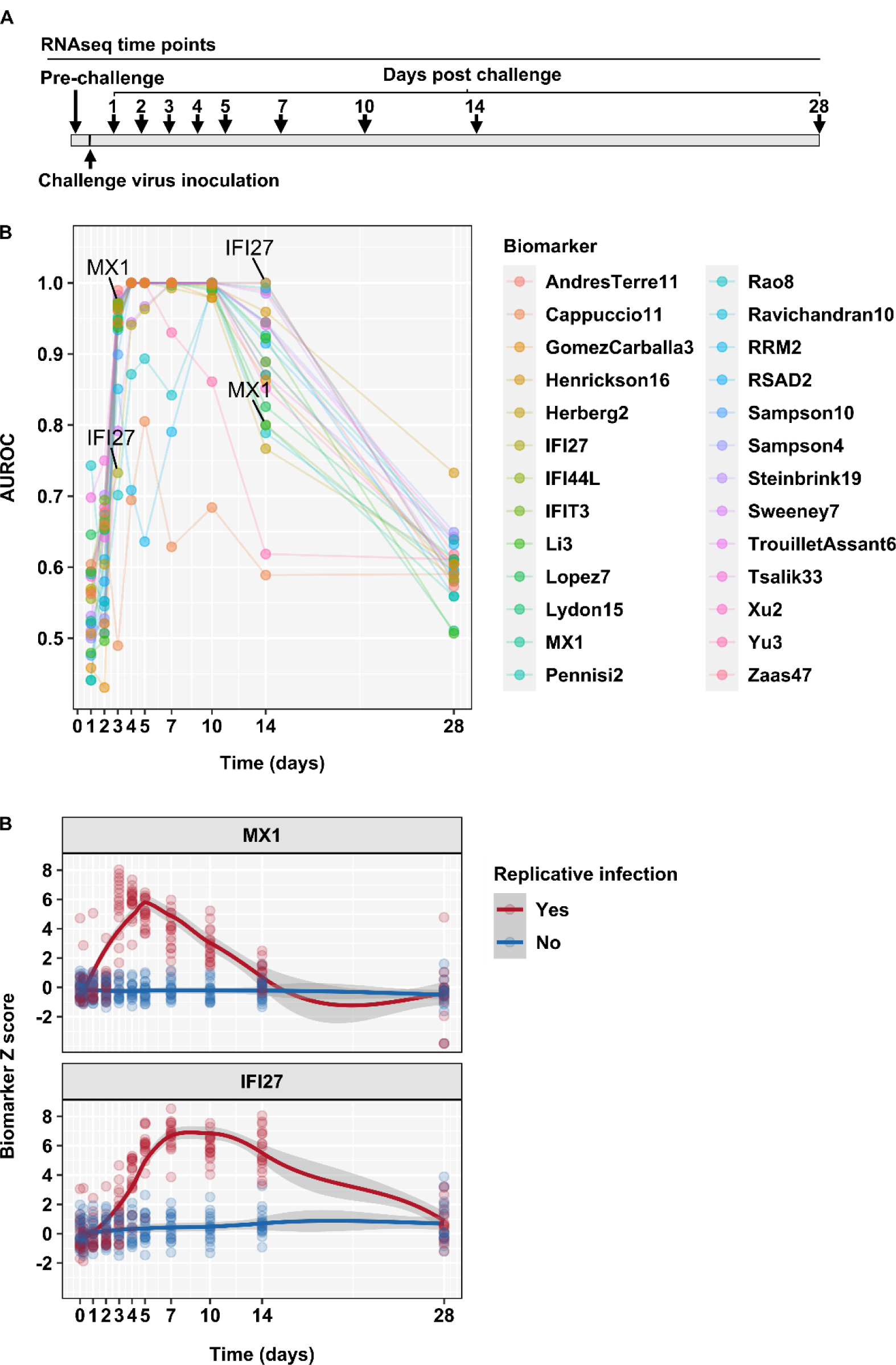
Blood transcriptional discrimination of participants with and without replicative infection by time from SARS-CoV-2 challenge. **(A)** Time points for blood RNA sampling in relation to virus challenge. **(B)** Point estimates for area under the receiver operating characteristic curve (AUROC) stratified by blood transcriptional signature and time after virus challenge. **(C)** Individual (data points) and loess smoothed summary (line ±95% CI) for standardised blood transcript levels of MX1 and IFI27 in sequential time points after challenge, for participants with (N=18) and without (N=16) replicative viral infection.

### Identification of blood transcriptional biomarkers of early and late phases of SARS-CoV-2 infection

Next, we compared the AUROC of each signature stratified by time point. Most achieved near perfect discrimination of participants with and without replicative infection in days 4-10 (Supplementary Figure 4A). We found greater variation in performance of each signature before and after this time interval, suggesting differential ability to identify early and late phases of viral infection. To investigate this hypothesis further, we focused on the single gene transcripts with highest AUROC on day 3 (MX1) and on day 14 (IFI27). On day 3, MX1 achieved an AUROC of 0.97 (0.93-1) which reduced to 0.8 (0.64-0.96) by day 14. In contrast, IFI27 achieved an AUROC of 0.73 (0.56-0.91) on day 3, increasing to 1 by day 14 (Figure 2B). These findings reflected an early but transient increase in MX1 expression and a comparatively delayed but sustained increase in IFI27 expression (Figure 2C). A number of other single gene biomarkers (IFI44L, IFIT3 and RSAD2) were highly correlated to MX1 and distinct from IFI27 (Supplementary Figure 4B).

### Relationship of MX1 and IFI27 expression in blood to symptoms and SARS-CoV-2 viral load

Most biomarker discovery and validation has focused on naturally acquired symptomatic viral infection. We and others have shown that host response biomarkers are able to detect asymptomatic infection^2, 39^. Consistent with this, we found no correlation between blood transcriptional scores and prospective quantitation of daily symptom scores among individuals who developed replicative infection (Figure 3A). We found elevated biomarker scores (>Z2 threshold) at time points in which participants who experienced replicative infection were completely asymptomatic. This was more evident with MX1 measurements at early time points and with IFI27 measurements at late time points.

**Figure 3.**
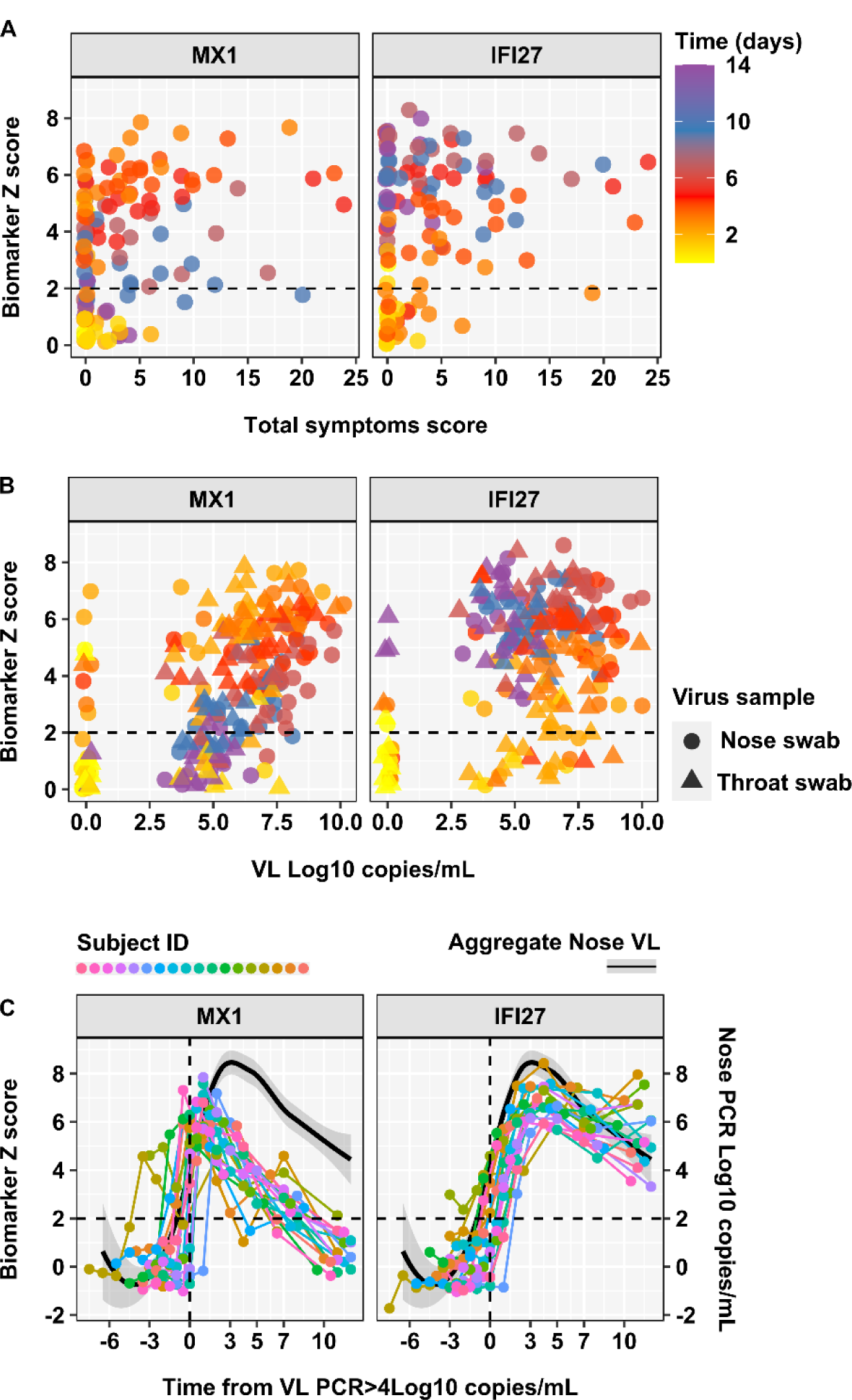
Relationship between blood transcript levels of MX1 and IFI27 with symptoms and viral load by time from SARS-CoV-2 challenge. Individual standardised blood transcript levels of MX1 and IFI27 against **(A)** symptom scores and **(B)** nose and throat viral loads in participants who developed replicative virus infection (N=18), stratified by time after virus challenge, with dashed line to represent the threshold (Z>2) for elevated transcript levels. **(C)** Individual standardised blood transcript levels of MX1 and IFI27 (connected data points, left axis) and loess smoothed summary for nose viral load (black line ±95% CI, right axis) in participants who developed replicative virus infection (N=18), by time from virus detection in nose swabs >4 Log10 copies/mL.

In addition, we investigated the relationship between blood transcriptional signature scores and viral load stratified by time from inoculation in samples from individuals who developed replicative infection. Examples of elevated MX1 and IFI27 scores (Z>2) were evident at time points with negative virus PCR in contemporary nose or throat swabs (Figure 3B). Elevated MX1 scores associated with negative virus PCR tests were more evident at early time points, and elevated IFI27 scores associated with negative virus PCR tests were more evident at late time points. Importantly, MX1 and IFI27 scores in the normal range (<Z2) were also evident at time points with positive virus PCR tests in contemporary samples. False-negative biomarker results were more evident for IFI27 at early time points, and for MX1 at late time points. To underscore the differential temporal relationship of each biomarker with viral load, we examined longitudinal biomarker measurements per participant who developed replicative infection, indexed by time from first PCR detection of virus (>4 Log10 copies/mL) in nasal swabs, which we have recently reported to correlate best with viral emissions^57^. The rise in MX1 scores was generally co-incident with PCR detection of the virus, and in some individuals evident before detection of virus by PCR. However, the MX1 response generally peaked before the peak in viral load, suggesting that clearance of MX1 transcript enrichment was faster than clearance of the virus. In contrast, IFI27 scores increased after detection of the virus and remained elevated after viral load started to fall (Figure 3C).

Both blood transcriptional biomarkers showed statistically significant correlation with viral load when including PCR negative time points (Supplementary Figure 5A) consistent with the fact that they provided good discrimination of groups of participants with and without replicative infection. However, when restricting the analysis to time points with positive virus PCR tests, we found a significant correlation only to MX1, suggesting this biomarker provided better prediction of viral load than IFI27 (Supplementary Figure 5B). Consistent with this observation, we also found that MX1 provided a better biomarker of infectiousness than IFI27, by predicting positive viral culture in contemporary samples. Among individuals who developed replicative infection, blood MX1 transcript levels discriminated virus culture positivity in nose or throat samples with AUROC 0.85 (0.79-0.92), significantly better than IFI27 which achieved AUROC of 0.66 (0.57-0.75). In this analysis false positive MX1 levels were limited to early time points, consistent with the observation that the rise in MX1 levels can precede PCR detection of the virus (Figure 4).

**Figure 4.**
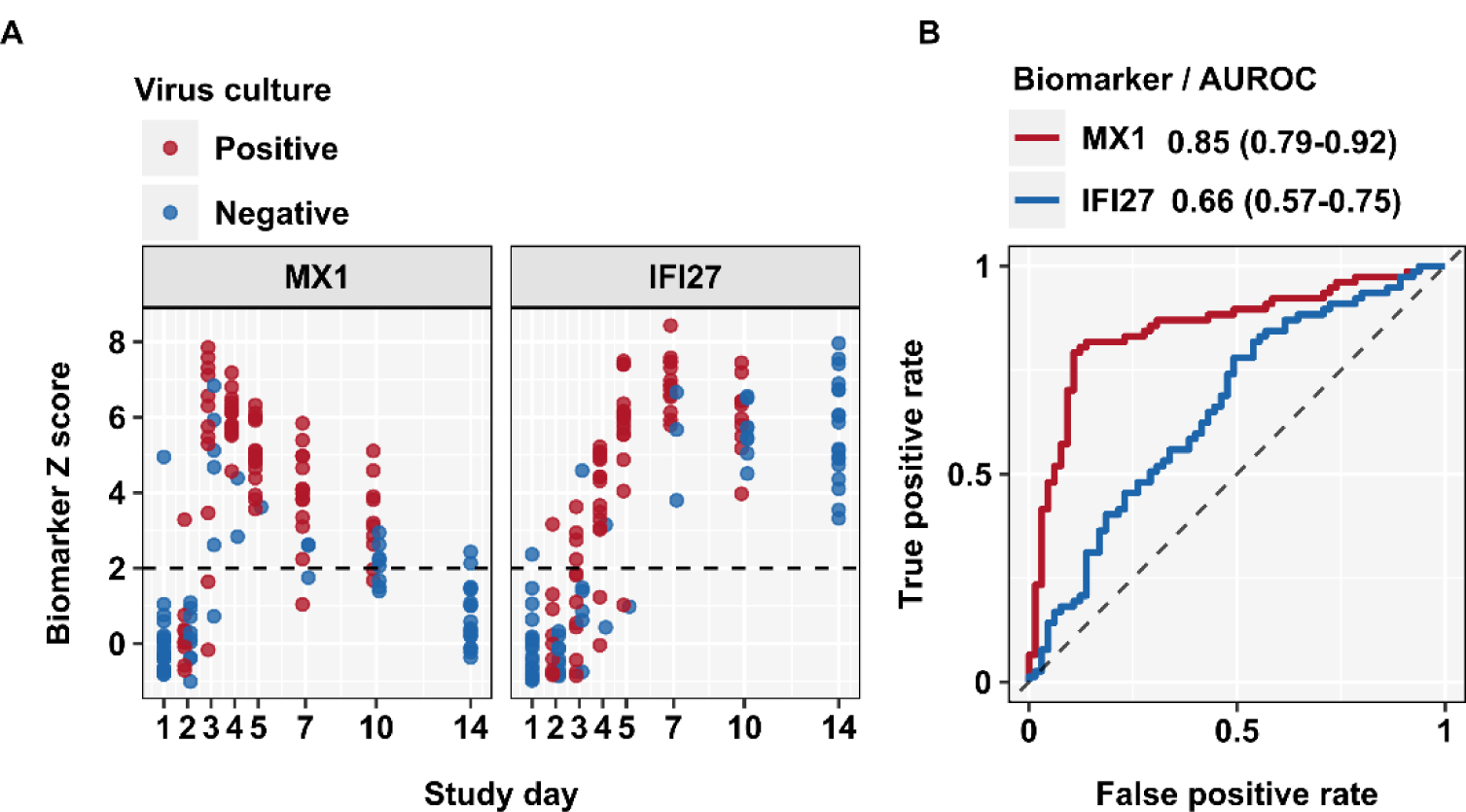
Discrimination of virus culture positivity by blood transcript levels of MX1 and IFI27. **(A)** Individual standardised blood transcript levels of MX1 and IFI27 at each time point for all individuals who develop replicative infection (N=18), stratified by contemporary virus culture positivity in either nose or throat swabs, with dashed line to represent the threshold (Z>2) for elevated transcript levels. **(B)** Area under the receiver operating characteristic curve (AUROC) discrimination of virus culture positivity by blood transcript levels of MX1 and IFI27 across all time points, showing AUROC point estimates and 95% confidence intervals.

### Differential regulation of MX1 and IFI27 expression in blood

Both MX1 and IFI27 are widely recognised as interferon stimulated genes (ISG)^58, 59^. To explore this relationship among participants in the replicative infection group, we compared MX1 and IFI27 levels with the average expression of a multigene signature (“STAT1 regulated module”) that we had previously derived and validated as a measure of type 1 IFN bioactivity^60^. Both biomarkers showed a statistically significant correlation with the STAT1 module, but the relationship with MX1 was stronger with near perfect correlation and covariance, suggesting that IFI27 expression was subject to additional levels of transcriptional regulation (Figure 5A-B). To obtain a deeper insight into the mechanisms of differential regulation of MX1 and IFI27, we investigated their expression in our previously reported single cell RNA sequencing analysis of PBMC from a subset of participants with replicative infection in the present SARS-CoV-2 challenge study^61^. We found a clear increase of MX1 expression in all major PBMC subsets in pooled day 3 data, and subsequent reduction by day 7. In contrast, increased expression of IFI27 was almost exclusively restricted to myeloid cells (monocytes and conventional dendritic cells). Modest upregulation was evident at day 3, but then increased further at day 7 and day 10 before reducing again by day 14, although expression levels remained higher than baseline through to day 28 (Figure 5C).

**Figure 5.**
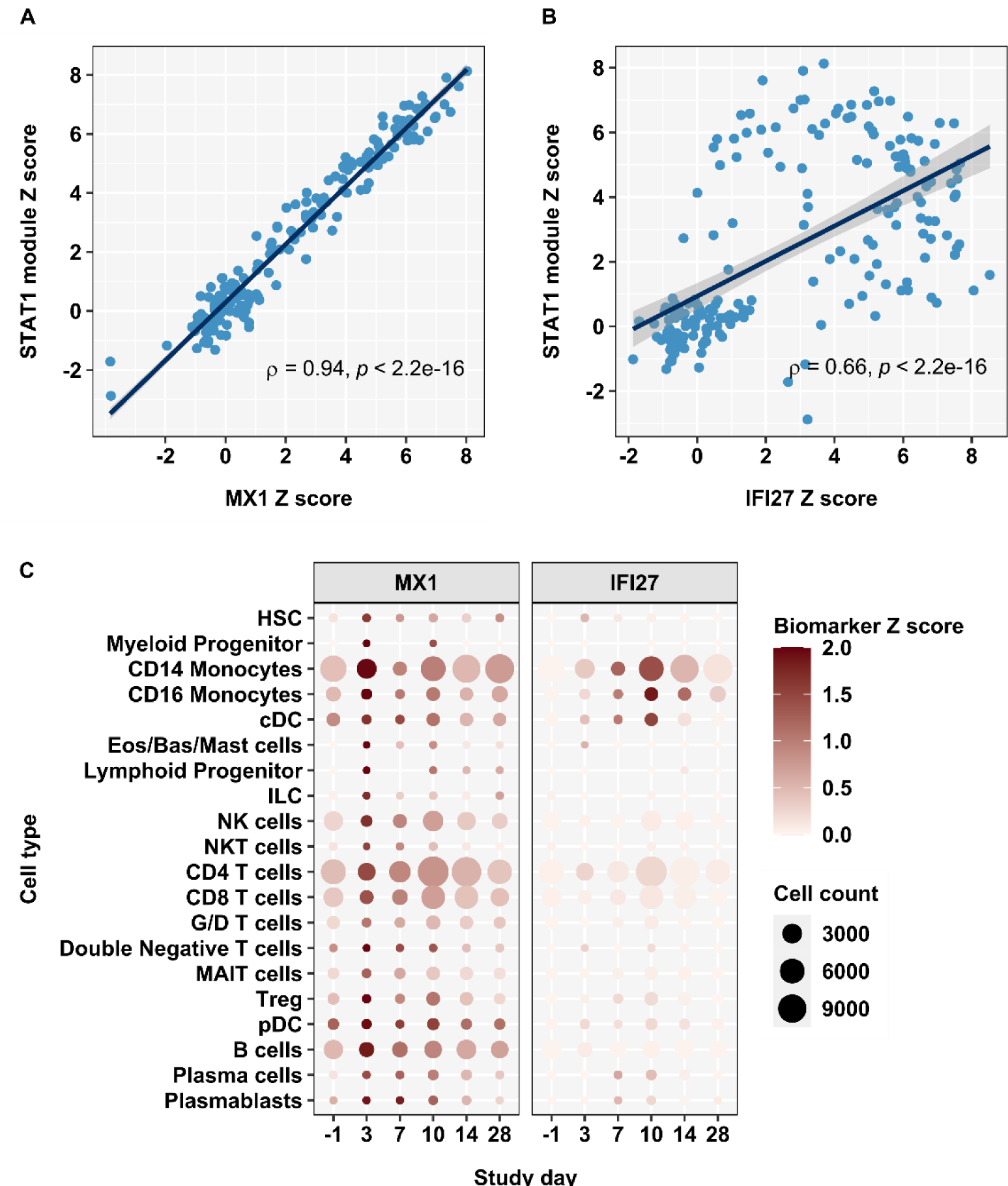
Differential regulation of MX1 and IFI27 expression in blood. **(A)** Individual standardised blood transcriptional expression of a type 1 IFN stimulated gene signature (STAT1 module) by standardised blood transcriptional expression of MX1 and IFI27 at all time points in participants who developed replicative virus infection (N=18), showing Spearmen correlation coefficients and p value. **(B)** Standardised blood transcriptional expression of MX1 and IFI27 stratified by cell type and time after virus challenge, in pooled data from participants who developed replicative virus infection (N=6).

In published ATAC sequencing data^20^, we tested the hypothesis that differential time and cellular distribution of MX1 and IFI27 expression reflected differential epigenetic regulation (chromatin accessibility) of *MX1* and *IFI27* loci in circulating immune cells. In datasets from unstimulated monocytes, CD4 T effector cells and B-cells from healthy individuals, we found evidence that the *MX1* locus contained areas of open chromatin (enrichment of sequencing peaks) close to the transcription start site and exon-1 (Supplementary Figure 6A-B), which would enable rapid transcriptional upregulation of this gene across multiple cell types. In contrast, the *IFI27* locus contained little evidence of open chromatin (Supplementary Figure 6A-B) in any of these cell types, and therefore inaccessible for rapid transcriptional upregulation. To evaluate subsequent epigenetic modifications following infection, we leveraged single cell ATACseq data from patients admitted to hospital with COVID-19^30^. Despite the sparsity in single cell data and relatively low coverage of the IFI27 locus, in samples from patients with acute COVID-19, we found a higher number of IF27 sequencing reads in monocytes compared to all lymphocyte populations. This difference was less evident in data from convalescent patients (Supplementary Figure 6C), and consistent with transient cell-type specific opening of the *IFI27* locus in established infection, providing a mechanistic basis for the temporal delay and cellular restriction of IFI27 responses compared to MX1.

### Generalisable differential utility of blood MX1 and IFI27 transcriptional biomarkers in acute respiratory virus infection

In order to investigate whether the differential host responses represented by MX1 and IFI27 were generalisable to other acute respiratory viral infections, we investigated their expression profiles in collated data from previously reported influenza, respiratory syncytial virus, and rhinovirus human challenges among participants with evidence of infection following inoculation as per original study definitions^17^. In every case MX1 upregulation in whole blood transcriptional profiles preceded that of IFI27 (Figure 6A). The data from these experiments were limited to approximately 6 days post-challenge and did not allow us to fully compare the temporal profiles of these biomarker measurements to the present SARS-CoV-2 challenge. Therefore, we undertook transcriptional profiling of blood samples from another recent H3N2 influenza human challenge model that included sampling beyond day 7^6^. This analysis also reproduced our findings in the SARS-CoV-2 challenge (Figure 6B).

**Figure 6.**
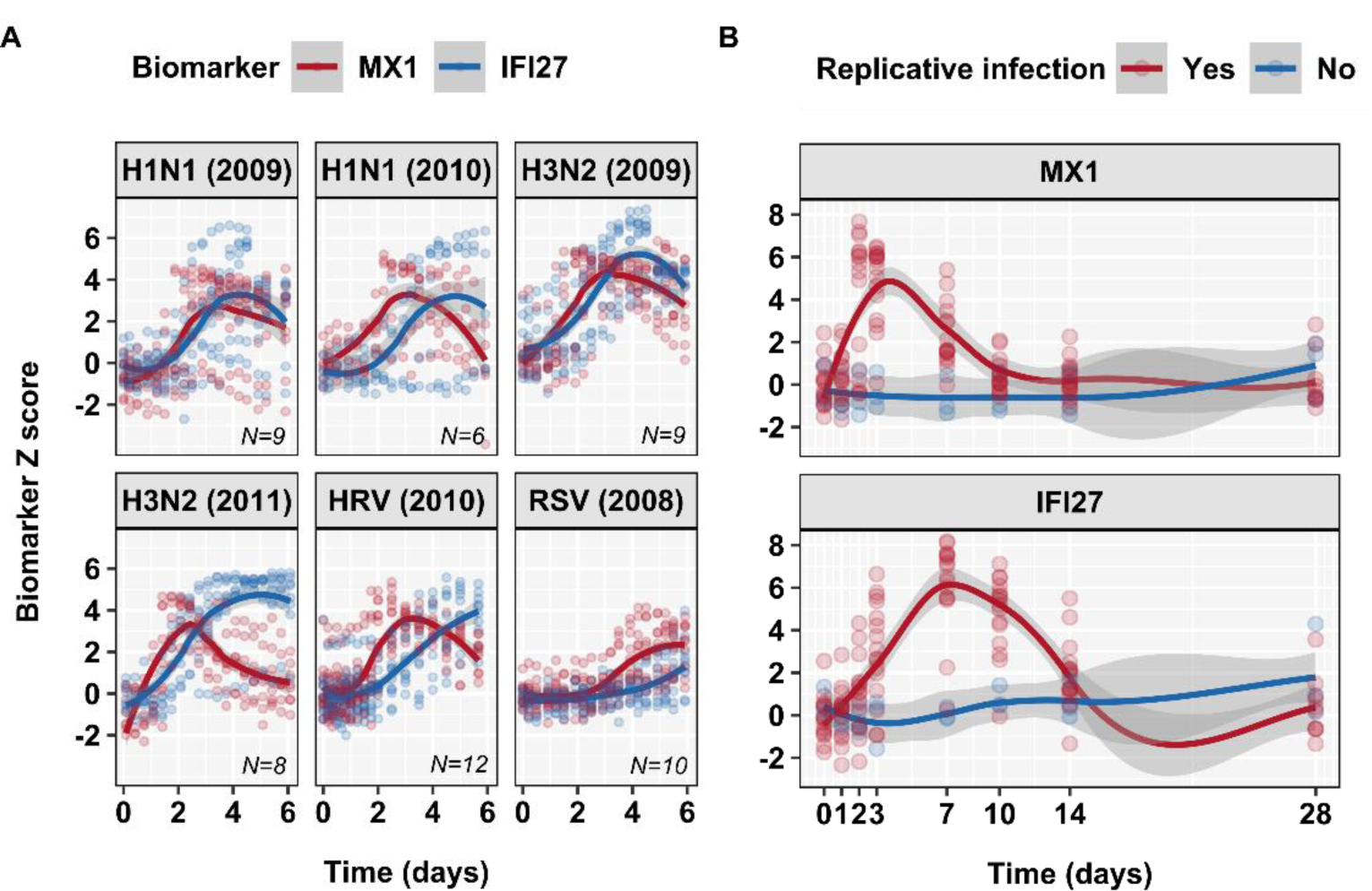
Generalisable differences in temporal profiles of blood MX1 and IFI27 expression in diverse respiratory virus challenges. **(A**) Individual (data points) and loess smoothed summary (line ±95% CI) for standardised blood transcript levels of MX1 and IFI27 over the first 6 days after challenge in selected human respiratory virus challenge models (GSE73072) among participants who develop replicative infection, and **(B)** over 14 days in an H3N2 influenza human challenge model, among participants with (N=16) and without (N=3) replicative infection.

We further sought to extend the generalisability of our findings to natural infections. In a household contact study of index cases with COVID-19^5^, blood transcript levels of MX1 and IFI27 achieved equivalently good discrimination of contacts with and without prevalent SARS-CoV-2 infection at recruitment (day 0, AUROC 0.97, 0.92-1). This level of discrimination was maintained for IFI27 in follow up samples 7 days later, but significantly reduced for MX1, consistent with earlier resolution of this biomarker (Figure 7A). In a further data set from patients with unselected community acquired respiratory virus infections, we evaluated MX1 and IFI27 expression in whole blood transcriptional profiles of individuals with PCR confirmed respiratory virus infections within 48 hours of symptom onset, in four sequential samples on alternate days^18^. Compared to baseline (pre-infection) samples from the same individuals, increased levels of MX1 expression (Z>2) were largely confined to early time points day 0-2 after presentation within 4 days of symptom onset. Increased levels of IFI27 expression (Z>2) were evident over a longer time course including day 4-6 after presentation, up to 8 days after symptom onset (Figure 7B, Supplementary Figure 7A). Across all time points, IFI27 measurements achieved statistically better AUROC than MX1 measurements for discrimination of infection from baseline uninfected samples (Supplementary Figure 7B). However, when the analysis was stratified by sample time point, MX1 achieved the highest AUROC for discrimination of infected samples on the day of presentation (Supplementary Figure 7C). The AUROC for MX1 reduced significantly at each subsequent time point. The time point stratified analysis of IFI27, showed stable AUROC discrimination of infection. A combined biomarker signature, comprising the average expression of MX1 and IFI27 improved the AUROC discrimination at early time points compared to IFI27 alone, and at late time points compared to MX1 alone (Supplementary Figure 7C).

**Figure 7.**
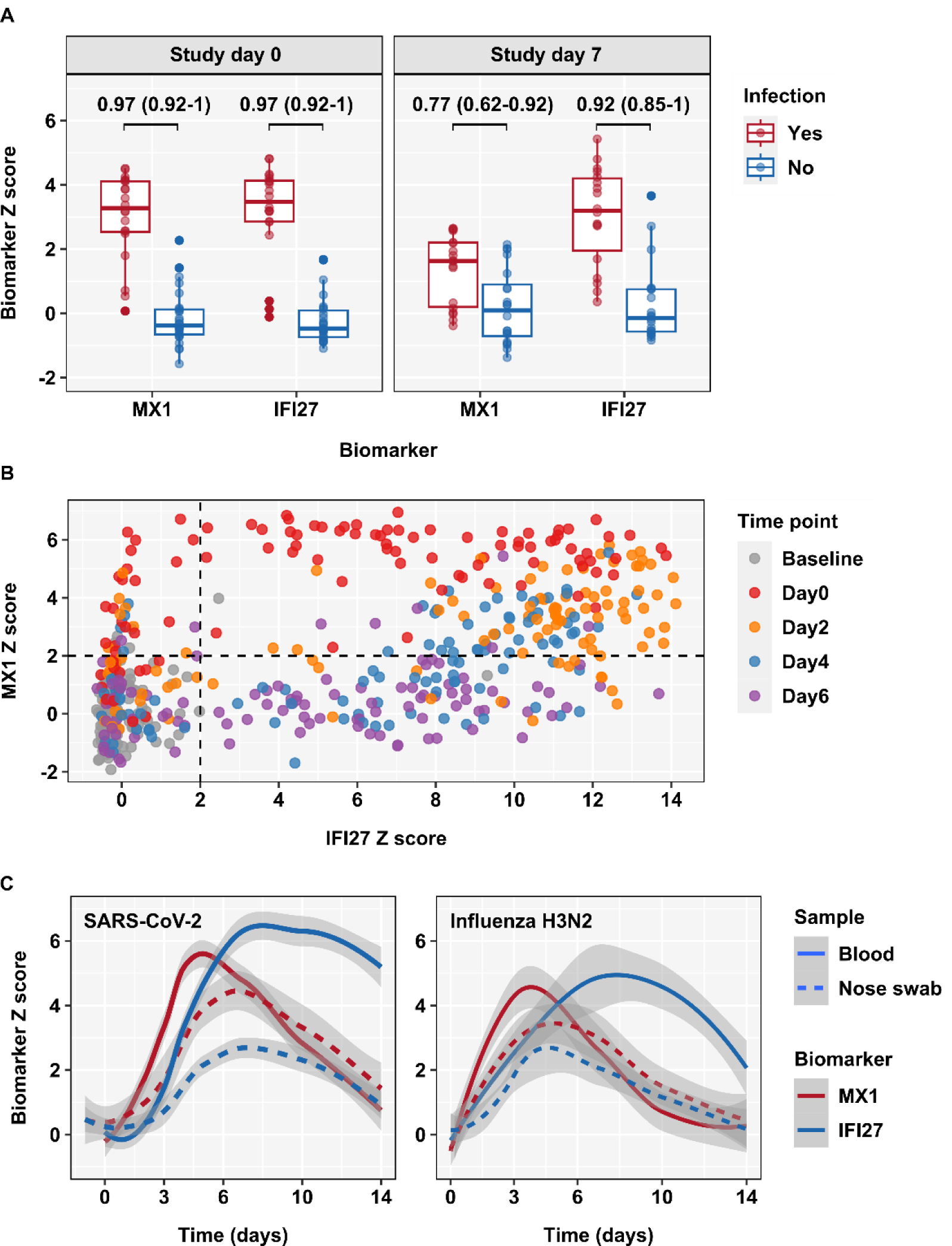
Differences in temporal profiles of blood MX1 and IFI27 expression in naturally acquired respiratory virus infections, and delayed responses in the nose to virus challenge. **(A)** Discrimination between SARS-CoV-2 infected (N=20) and uninfected (N=26) household contacts of index cases with COVID-19 by blood transcript levels of MX1 and IFI27, at participant recruitment (study day 0) and 7 days later. Data points represent individual study participants, summarised by box and whisker plots. Discrimination accuracy is shown as AUROC point estimate and 95% confidence intervals. **(B)** Individual standardised blood transcript levels of MX1 against IFI27 for sequential samples before infection (baseline, N=128) and at time points indicated (day0, N=103; day 2, N=106, day 4, N=100; day 6, N=102) after presentation within 48 hours of symptoms onset among prospectively recruited participants with unselected respiratory virus infections. **(C)** Loess smoothed summary (line ±95% CI) for standardised transcript levels of MX1 and IFI27 in blood (N=18) and nose samples (N=5-13) from participants who developed replicative virus infection by time after SARS-CoV-2 challenge, and in blood (N=16) and nose samples (N=12-13) from participants who developed replicative virus infection by time after H3N2 influenza challenge.

### Comparison of host response biomarkers of acute respiratory virus infection in blood and nose samples

The potential to measure host response transcriptional signatures in samples from upper respiratory tract swabs has recently been reported^62, 63^. We compared MX1 and IFI27 transcript measurements in samples from blood and nose swabs in the present SARS-CoV-2 challenge. Surface nose swabs only yielded adequate RNA for sequencing in 103 of 238 samples (43%), reflecting an inherent technical limitation in this approach. Nonetheless, for nose samples which did yield RNA sequencing data, we found clear evidence of MX1 and IFI27 responses in participants who developed a replicative infection. In comparison to blood measurements of these biomarkers, the signal strength in nose swab samples was weaker than in blood, the response in the nose was delayed in comparison to blood, and the differential time course for each biomarker evident in blood samples was lost in nose swab samples (Figure 7C). These findings were replicated in blood and nasal mucosal curettage samples from the H3N2 influenza human challenge and indicate that in general, blood biomarker measurements are likely to provide better diagnostic discrimination for prevalent infection as well as better differentiation of early and late phases of infection, compared to nasal swabs (Figure 7C).

## Discussion

We present a comprehensive evaluation of previously reported transcriptional signatures as host response biomarkers of viral infection in high frequency longitudinal blood and nose swab samples from the first SARS-CoV-2 human challenge experiment. We provide compelling evidence showing that single gene transcripts for MX1 and IFI27 in blood, discriminate temporally distinct phases of infection, and we show that these findings are generalisable across a range of clinically important respiratory viruses in both experimental and naturally acquired infections. The earliest phase of replicative SARS-CoV-2 infection was associated with rapid upregulation of MX1 transcripts in blood, which may precede PCR detection of the virus and correlated with PCR positive viral load measurements. In contrast, blood transcriptional upregulation of IFI27 occurred after PCR detection of the virus. IFI27 expression did not correlate with PCR positive viral load measurements and was sustained above baseline levels after viral clearance. Of note, transcriptional upregulation of both biomarkers was independent of symptoms.

Both MX1 and IFI27 are widely recognised as ISGs^58, 59^. The MX1 response closely reflected generalised type 1 ISG expression across all major cell types. We focused on MX1 because it achieved the highest single gene point estimate AUROC for discriminating groups of individuals with and without replicative viral infection at the first time point at which any biomarker achieved significant discrimination. Alternative interferon inducible single gene biomarkers such as IFI44L, IFIT3 and RSAD2 provided statistically comparable discrimination at this time point, and are highly correlated to MX1. These biomarkers are likely to share the same mechanisms for transcriptional regulation, and offer the same utility as MX1. Delayed transcriptional upregulation of IFI27 compared to other canonical ISGs has also been reported following in vitro stimulation of cells with IFN^64, 65^. In vivo, IFI27 expression in blood samples was restricted to myeloid PBMC. We found evidence of differential epigenetic silencing of the IFI27 locus compared to the MX1 locus in resting PBMC, and cell type specific epigenetic modulation of this locus in monocytes during established COVID-19 infection. These data provide a mechanistic explanation for the differential temporal and cellular expression of the two biomarkers, namely that IFI27 is epigenetically silenced in resting cells but becomes accessible for transcription in specific myeloid lineages during the evolving immune response to infection.

The differential temporal expression of MX1 and IFI27 in the SARS-CoV-2 challenge model was replicated in challenge experiments with multiple influenza strains, respiratory syncytial virus, or rhinovirus, and in data from household contacts with naturally acquired SARS-CoV-2 infection. Likewise, in unselected community acquired symptomatic respiratory virus infections, in which MX1 measurements achieved high diagnostic accuracy for infections within 4 days of symptom onset, and IFI27 measurements achieved higher diagnostic accuracy at later time points. The combination of both measurements provided highest diagnostic accuracy across all time points.

In SARS-CoV-2 and H3N2 Influenza challenge experiments, we found no evidence that measuring these biomarkers in nose swab samples offered any advantage to blood samples. Lower upregulation of gene expression in nose samples compared to blood, and the loss of temporal differentiation between the biomarkers may reflect the biology of superficial cell populations compared to circulating leukocytes. Unexpectedly, upregulation of these biomarkers in nose samples of individuals with replicative infection was also temporally delayed compared to the blood. This finding is also evident in our analysis of comparative single cell sequencing data from blood and nose swab samples^61^. Whether it reflects faster transmission of IFN signalling to circulating blood cells, later onset of viral replication in the nose compared to the throat, or local suppression of IFN signalling by the virus in the nasal mucosa require future mechanistic investigation.

We propose that these blood transcriptional biomarkers of early and late phase of viral infection offer a range of research and clinical applications. Upregulation of MX1 expression may be used to detect pre-symptomatic infection in contacts of index cases. Its correlation with viral load and culture results suggests it may also provide clinical utility to infer infectiousness and thus trigger infection control interventions. Finally, since specific antiviral treatment efficacy diminishes with duration of infection^66, 67^, MX1 measurements may be used to stratify patients most likely to benefit from treatment. Conversely, IFI27 expression may offer a more time-stable diagnostic triage test for viral infection due to sustained upregulation beyond the initial acute phase of infection. To account for differential temporal profiles, a combined approach including both MX1 and IFI27 (as averaged expression, or where a positive test for either gene triggers further confirmatory testing) may be the optimal approach to diagnostic triage.

Comprehensive identification of reported blood transcriptional biomarkers of viral infection by systematic review, and their application in standardised human SARS-CoV-2 and influenza human challenges with high frequency sampling are major strengths of this study, thus enabling identification of differential temporal profiles of MX1 and IFI27 responses. Single cell data from the SARS-CoV-2 challenge model, and analyses of publicly available data also allowed investigation of the mechanism for differential temporal profiles of MX1 and IFI27 responses, and to confirm reproducibility of our findings across a range of respiratory virus infections.

Our conclusions are currently limited to data derived from individuals with non-severe infection. Therefore, future validation in hospitalised cohorts for whom the time of exposure can be estimated will be required to assess whether severe disease alters the temporal profiles of these biomarkers. Finally, we do not address the specificity of our findings for respiratory virus infections. Therefore, we have limited our discussion of potential translational applications to diagnostic triage tests to trigger confirmatory virological investigations, stratification of patients with confirmed viral infections for antiviral treatment, and pre-symptomatic screening of contacts of index cases of confirmed viral infections. Notably, for each of these applications, the generalisability of blood transcriptional biomarkers across respiratory viruses may be considered a strength.

Translation of these viral biomarkers to near-patient platforms is now required to enable further evaluation of clinical utility and impact in prospective observational and interventional studies.

## Contributions

JR, RKG and MN conceived this study. RH, CC and MN obtained funding for this study. MK, AM, AC, BK, WB and CC conceived, obtained funding and supervised conduct of the SARS-CoV-2 challenge study. CT, LKB, RKG, JGB and MN undertook the systematic review of blood transcriptional biomarkers of viral infection. JR, RKG, LKB and MN undertook analysis of publicly available bulk RNAseq and ATACseq data. JR, CT, TM, LKB, JGB, HW, BL, CW, CV, BMC and RH undertook sample processing and data analysis for the SARS-CoV-2 challenge study. RL, LD, MZN and ST contributed single cell RNAseq data from the SARS-CoV-2 challenge study. CMB, LP, PD, MK, ML and CC contributed bulk data from the H3N2 influenza challenge study. KM, EC, JF, SH and AL contributed data from the INSTINCT SARS-CoV-2 household contact study. AJK and JCK contributed data single cell ATACseq data from the COMBAT study. JR, RKG and MN wrote the manuscript with input from all the authors.

## Funding acknowledgements

This research was supported by the Wellcome Trust (224530/Z/21/Z). AL acknowledges funding by the NIHR Health Protection Research Units (HPRU) in Respiratory Infections (NIHR200927). BMC acknowledges funding by the Rosetrees Foundation. CMB, MK and ML acknowledge funding by NIHR Biomedical Research to Imperial College London. CMW acknowledges funding from the Medical Research Council (MR/T016329/1). CT acknowledges funding from the Wellcome Trust (102186/B/13/Z). LCKB acknowledges funding from the NIHR (Academic Clinical Fellowship Programme). LMD acknowledges funding from the European Union’s Horizon 2020 research and innovation programme under the Marie Skłodowska-Curie grant agreement No 955321. M.Z.N. acknowledges funding from a MRC Clinician Scientist Fellowship (MR/W00111X/1), Action Medical Research (GN2911) and funding from the Rutherford Fund Fellowship allocated by the MRC UK Regenerative Medicine Platform 2 (MR/5005579/1). MN acknowledges funding from the Wellcome Trust (207511/Z/17/Z) and by NIHR Biomedical Research Funding to UCL and UCLH. R.H. is a NIHR Senior Investigator. RKG acknowledges funding from the National Institute for Health Research (NIHR302829).

## Data sharing statement

Processed RNAseq data are available from ArrayExpress from the SARS-CoV-2 challenge study (accession number: E-MTAB-12993), H3N2 influenza challenge study (accession numbers: E-MTAB-13038 and E-MTAB-13041). Raw sequencing data from these studies and the INSTINCT SARS-CoV-2 household contact study will be made available via the European Genome-Phenome Archive under managed data access.

## Supporting information

Supplementary File 1_SystematicReviewSummary

## Data Availability

Processed RNAseq data will be made available after peer reviewed publication at ArrayExpress for the SARS-CoV-2 challenge study (accession number: E-MTAB-12993), H3N2 influenza challenge study (accession numbers: E-MTAB-13038 and E-MTAB-13041). Raw sequencing data from these studies and the INSTINCT SARS-CoV-2 household contact study will be made available via the European Genome-Phenome Archive under managed data access.

