## Supplementary Figures for "SARS-CoV-2 human challenge reveals single-gene blood transcriptional biomarkers that discriminate early and late phases of acute respiratory viral infections"

***Supplementary Figure 1***

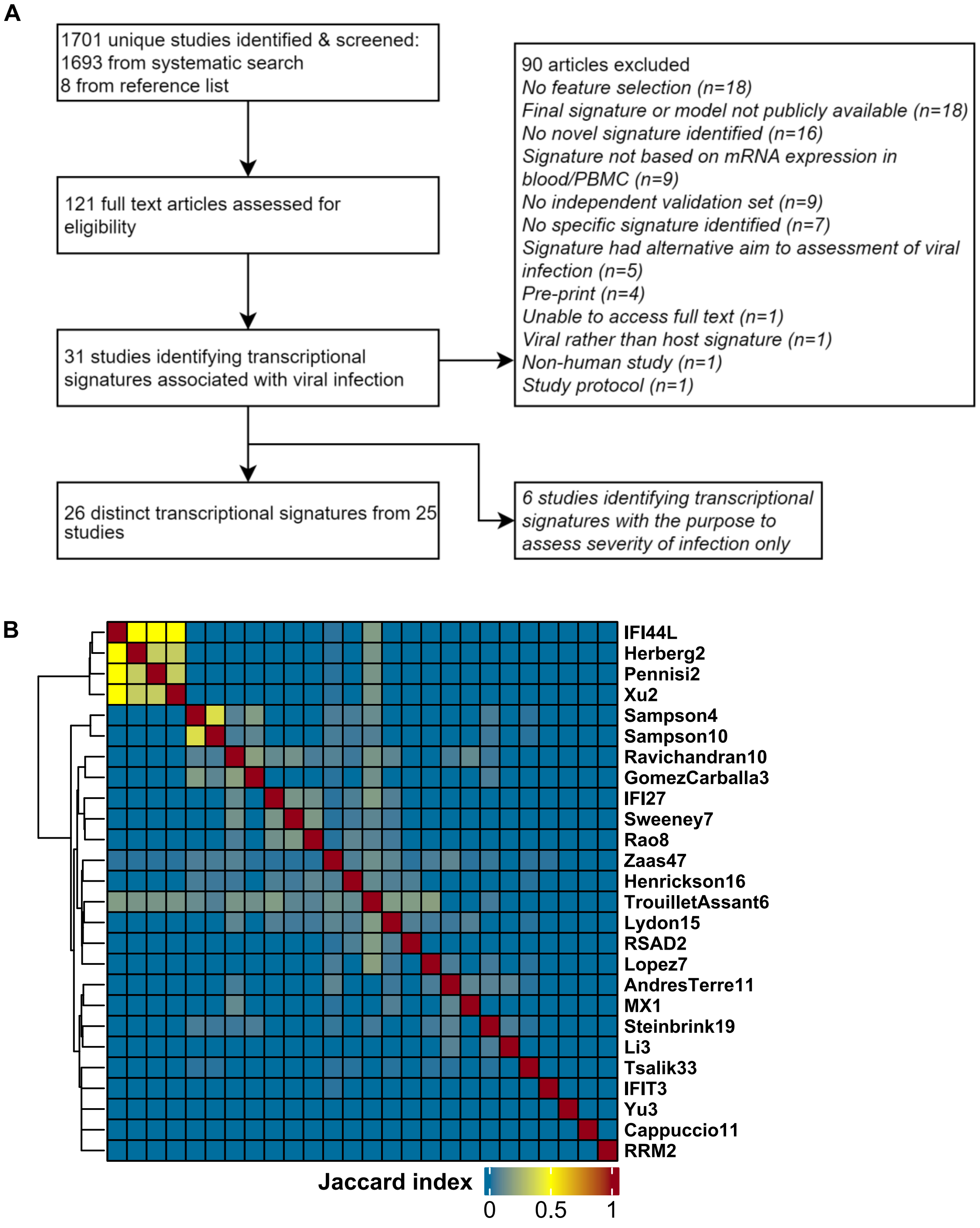

***Identification of blood transcriptional biomarkers of viral infection by systematic review and comparison of their gene composition.***

**(A)** Consort diagram of systematic review for identification of blood transcriptional biomarkers of viral infection. **(B)** Jaccard index pairwise comparisons of gene composition for each blood transcriptional signature in a symmetrical matrix with complete linkage clustering.

***Supplementary Figure 2***

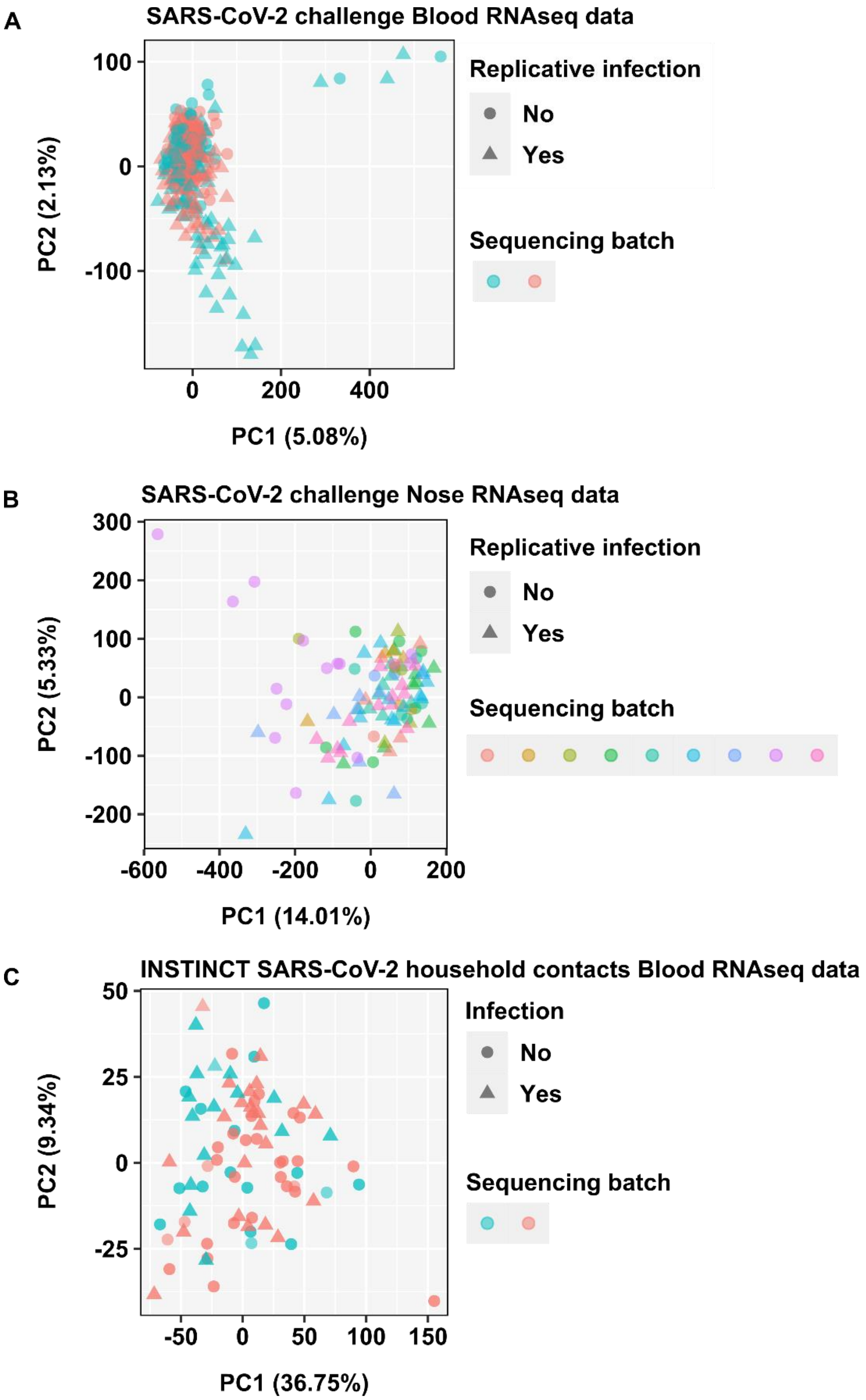

***Principal component analysis of RNA sequencing data***

Principal component analysis of **(A)** blood and **(B)** nose RNA sequencing data from the SARA-CoV-2 challenge model, and **(C)** blood RNA sequencing data from the INSTINCT SARS-CoV-2 household contacts study, stratified by replicative infection and/or sample processing batch.

***Supplementary Figure 3***

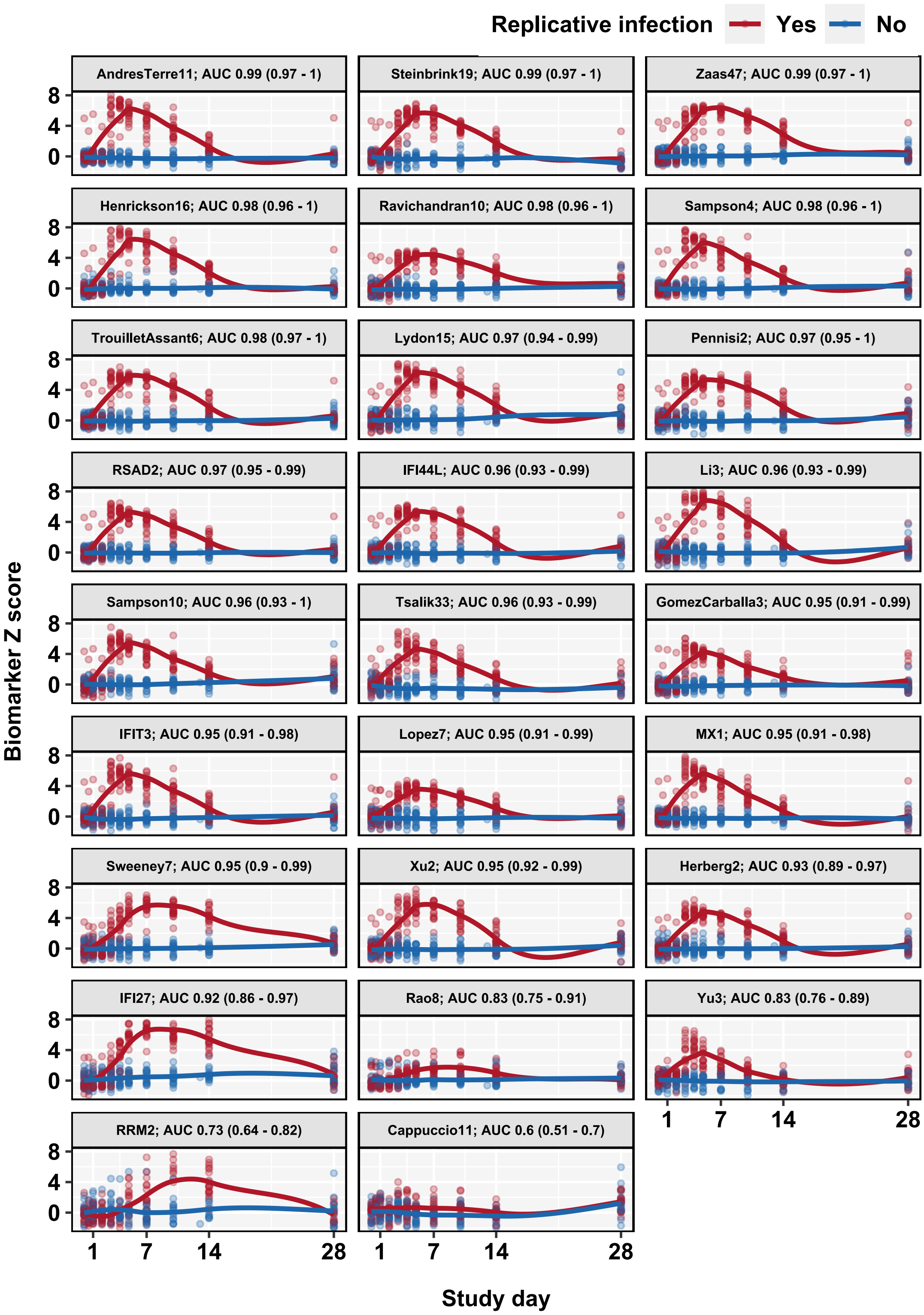

***Temporal profiles of blood transcriptional signature scores in participants with and without replicative SARS-CoV-2 infection.***

Individual (data points) and loess smoothed summary (line ±95% CI) for standardised blood transcript levels of each blood transcriptional signature in sequential time points after challenge, ranked in descending order of AUROC (±95% CI) for discrimination of participants with (N=18) and without (N=16) replicative viral infection using data from day 3, 7, 10 and 14.

***Supplementary Figure 4***

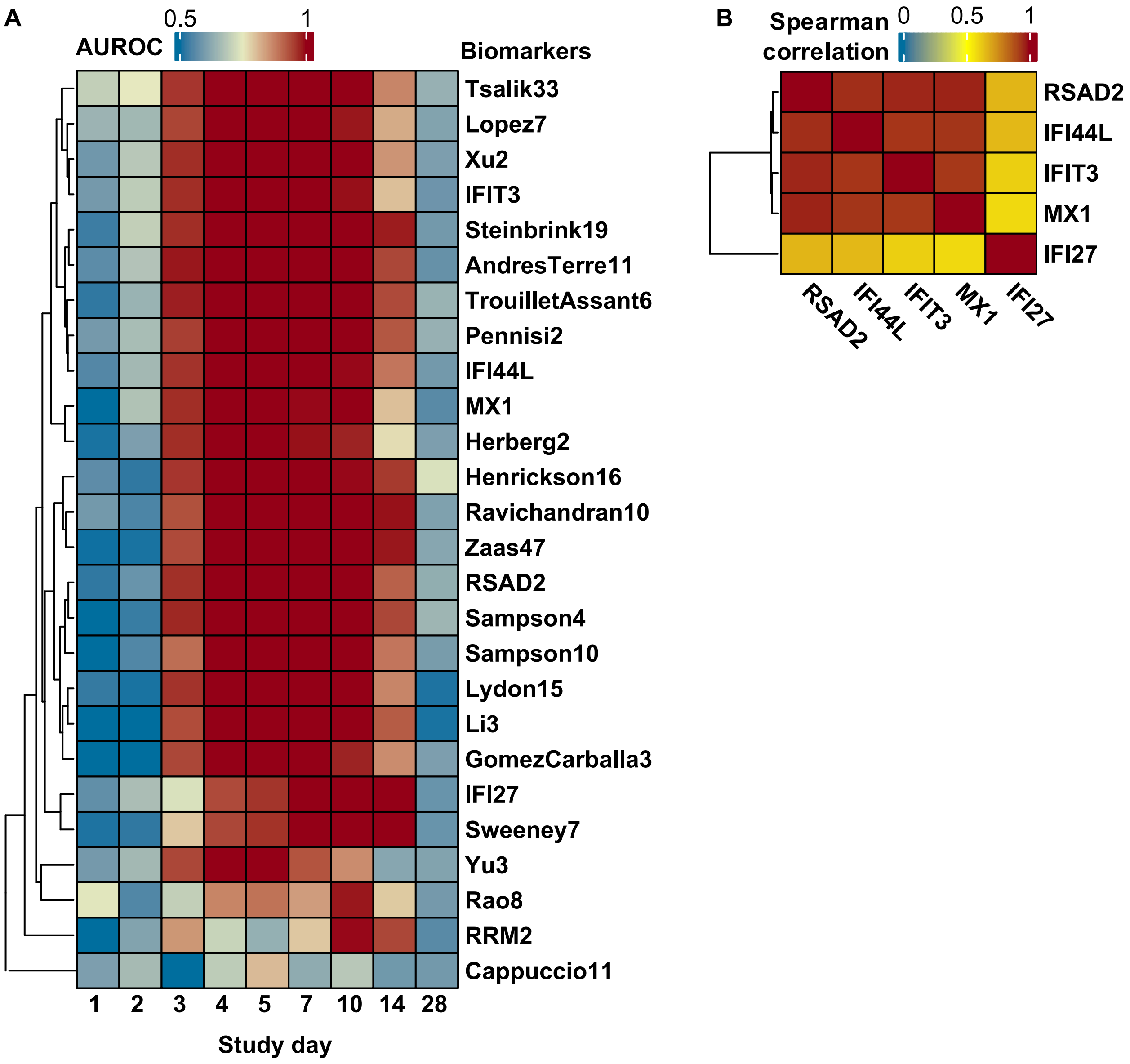

***Discrimination of participants with and without replicative SARS-CoV-2 infection by blood transcriptional biomarkers of viral infection.***

**(A)** Heatmap of AUROC point estimates for discrimination of participants with and without replicative SARS-CoV-2 infection by each blood transcriptional signature (rows) stratified by study day (columns). **(B)** Spearman correlation of selected interferon stimulated single gene blood transcriptional biomarker scores across all time points in participants with replicative SARS-CoV-2 infection (N=18).

***Supplementary Figure 5.***

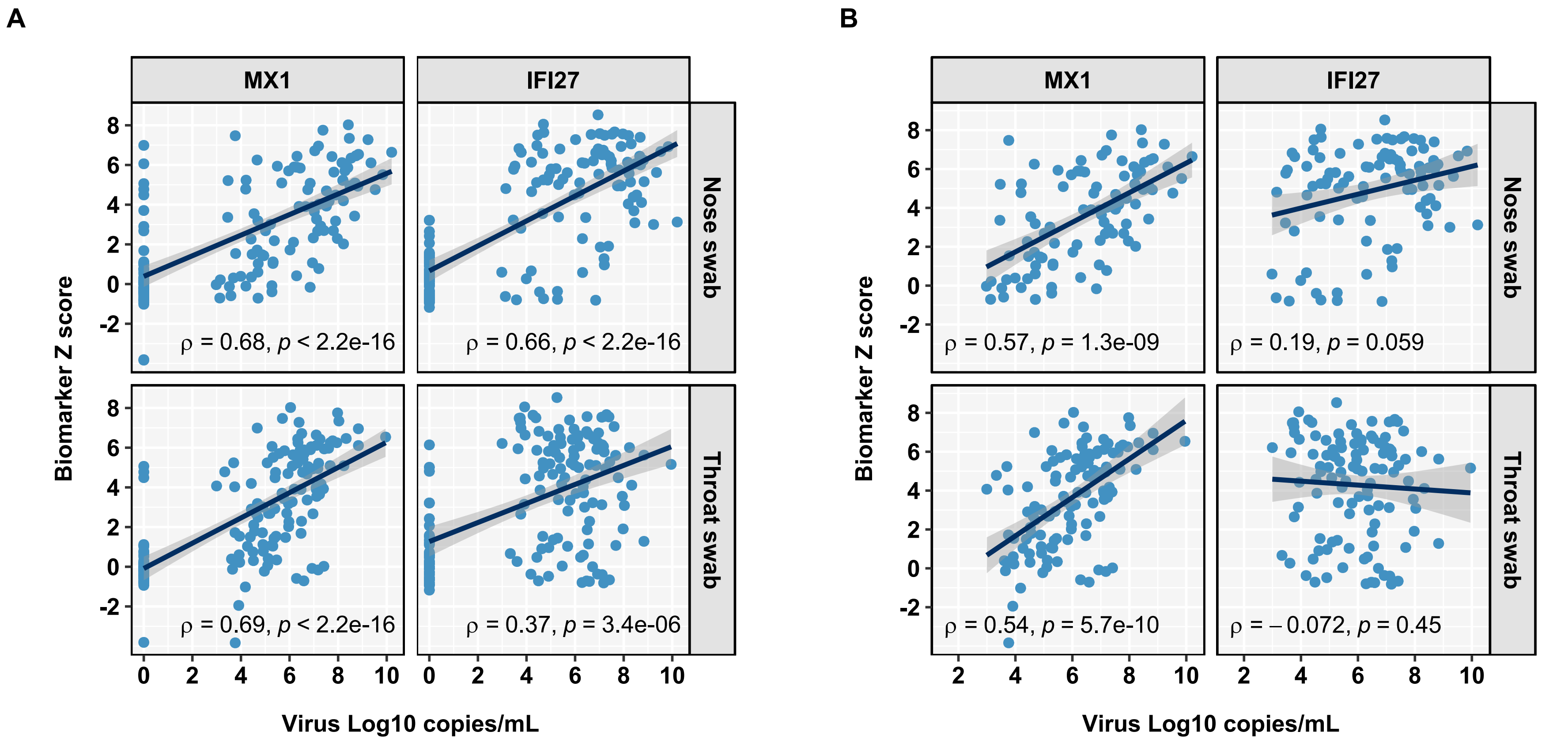

***Blood MX1 and IFI27 blood transcript correlation with PCR viral load measurements.***

Scatter plot and Spearman correlations of blood MX1 or IFI27 transcript levels with nose or throat PCR viral load measurements in participants with replicative SARS-CoV-2 infection (N=18) for all sampling time points **(A)** and for time points with detectable virus by PCR **(B)**.

***Supplementary Figure 6.***

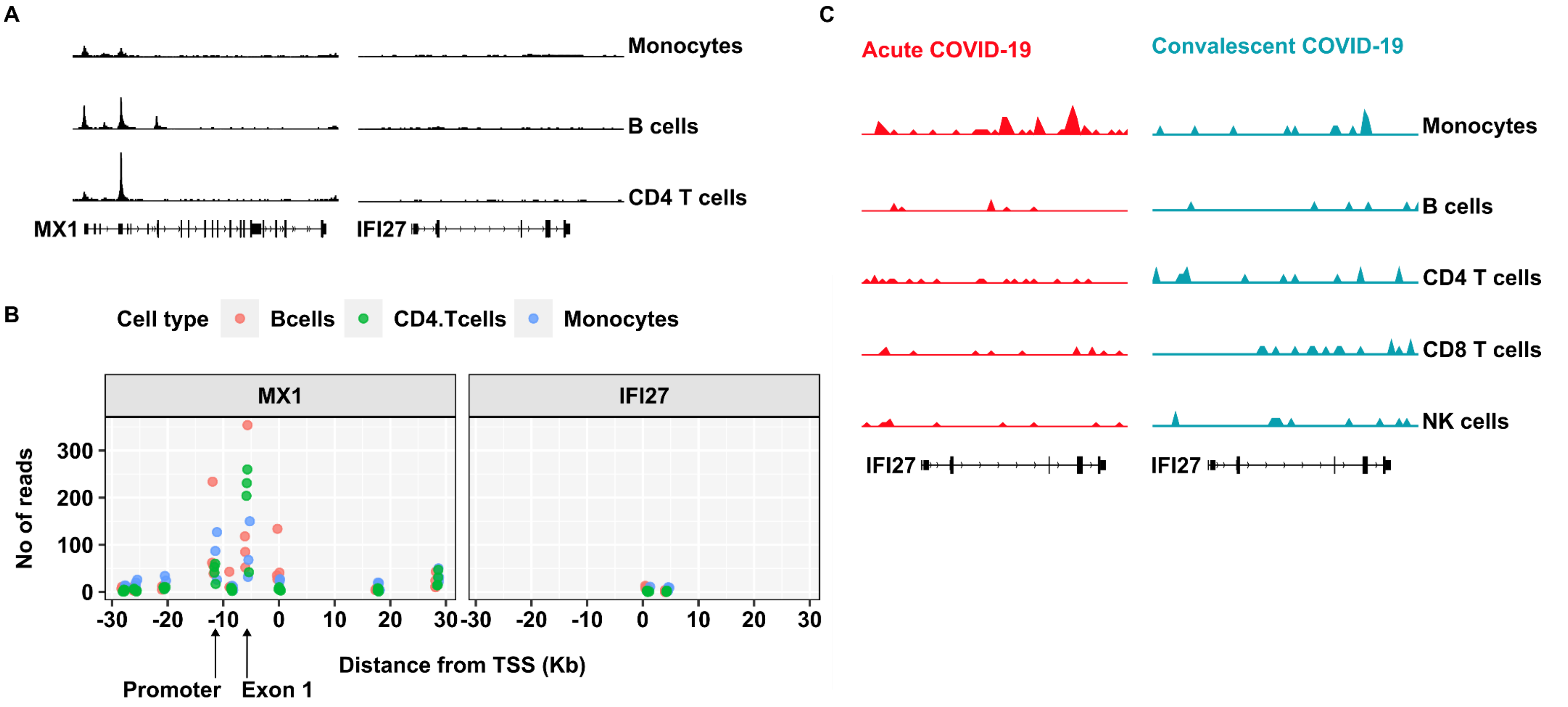

***ATAC sequencing reads at MX1 and IFI27 locus in unstimulated peripheral blood mononuclear cells.***

**(A)** Merged ATACseq read tracks (normalised ATAC signal range 0-1.8) from healthy donor sorted unstimulated monocytes (donor N=3), B-cells (N=4) and CD4 T effector cells (N=4) and **(B)** read counts are shown on the Y-axis, with genetic distance between the relevant peak and the gene's transcription start site (TSS) shown on the X-axis. (**C**) Merged single cell ATACseq read tracks at the *IFI27* locus (normalised ATAC signal range 0-0.04) for cell types indicated, in acute and convalescent samples from patients admitted to hospital with COVID-19 (N=8).

***Supplementary Figure 7.***

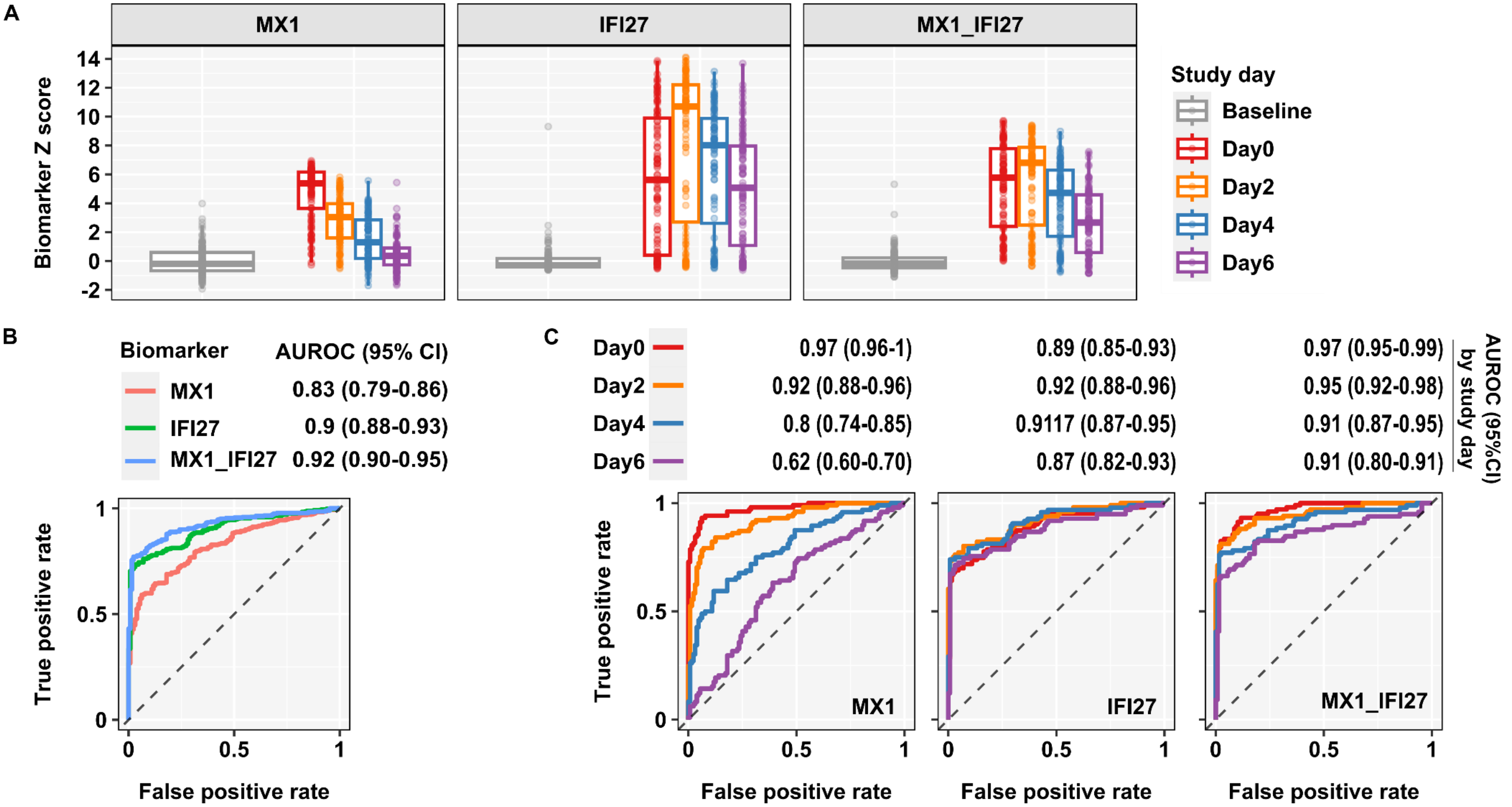

***Blood transcript discrimination of post infection time points from pre-infection samples in in unselected community acquired respiratory virus infections from GSE68310.***

**(A)** Individual data points and box plot summaries of blood transcript levels of MX1, IFI27 and average of MX1 and IFI27 in pre-infection baseline samples (N=128) and sequential time points on alternate days from study day 0 (up to 48 hours after onset of symptoms) in unselected community acquired respiratory virus infections (N=102-106). **(B)** Receiver operating curve discrimination of all post-infection samples from pre-infection baseline samples by blood transcript levels of MX1, IFI27 and average of MX1 and IFI27 with AUROC point estimates (±95% CI). **(C)** Receiver operating curve discrimination of post-infection samples stratified by study day from pre-infection baseline samples by blood transcript levels of MX1, IFI27 and average of MX1 and IFI27 with AUROC point estimates (±95% CI).
