## Supplementary Tables for "SARS-CoV-2 human challenge reveals single-gene blood transcriptional biomarkers that discriminate early and late phases of acute respiratory viral infections"

### *Supplementary Table 1: Characteristics of whole-blood RNA signatures for viral infection included in analysis*

| Signature(s) | Model[&] | Discovery population(s) | Discovery setting(s) | Discovery approach | Validation population(s) | Intended application |
|---|---|---|---|---|---|---|
| **AndresTerres11 (1)** | Geometric mean of all genes (influenza meta-signature) | Five cohorts of children and adults with influenza; adults challenged with influenza; and adults with bacterial pneumonia | UK, USA, and Australia | Differential expression followed by leave-one-cohort-out strategy and filtering for heterogeneity of effect size, using genome-wide data | Eight cohorts of children or adults with influenza or bacterial infection; adults challenged with influenza; and adults vaccinated against influenza | Influenza *vs* bacterial or other viral infection |
| **Cappuccio11(2)** | Difference in geometric means between upregulated and downregulated genes (COVID-19 Signature) | 84 publicly available transcriptomic studies classified into four categories: COVID-19 contrasts; other viral infection (respiratory and non-respiratory) contrasts; bacterial infection (respiratory and non-respiratory) contrasts; non-infectious contrasts. A typical contrast included samples from diseased subjects and healthy controls | Worldwide | Multi-objective fitness function that evaluates any proposed signature along three dimensions: detection, consistency with ATAC-seq and pathway annotation, and cross-reactivity. The multi-objective fitness function was optimized in training studies to return a population of high-fitness candidate signatures which were further selected based on performance in a set of development studies. The signature showing the most consistent performance in both training and development studies was selected. | 43 publicly available transcriptomic studies classified into four categories: COVID-19 contrasts; other viral infection (respiratory and non-respiratory) contrasts; bacterial infection (respiratory and non-respiratory) contrasts; non-infectious contrasts. A typical contrast included samples from diseased subjects and healthy controls | COVID-19 infection *vs* healthy, other bacterial/viral infections and non-infectious conditions |
| **Gómez-Carballa3 (3)** | Logistic regression§ | Aggregated data from 11 publicly available cohorts containing children and adults with viral or bacterial infections | USA, UK, Spain, The Netherlands, Australia, Mexico | Parallel Regularized Regression Model Search (4) on 64 candidate genes previously reported in the 11 cohorts to distinguish viral from bacterial infection | Split test cohort of independent samples from the aggregated dataset of 11 publicly available cohorts containing children and adults with viral or bacterial infections | Viral *vs* bacterial infection |
| **Henrickson16 (5)** | Difference in geometric means between upregulated and downregulated genes (influenza paediatric signature score) | Four cohorts of children with influenza-like illness | USA | Meta-analysis and leave-one-out strategy to identify common genes using genome-wide data | Two cohorts of children or adults with influenza | Influenza infection *vs* healthy |
| **Herberg2 (6)** | Sum of downregulated genes subtracted from the sum of upregulated genes (Disease Risk Score) | Children with viral or bacterial infection | UK, USA, and Spain | Elastic net followed by forward selection–partial least squares, using significantly differentially expressed transcripts | Children with bacterial or viral infection, inflammatory disease, or indeterminate diagnosis | Viral *vs* bacterial infection in febrile children |
| **IFI27 (7)[1]** | NA | Two cohorts of adults with influenza & one cohort of adults with | Australia, Canada, Germany | Differential gene expression between influenza cases and healthy control. IFI27 | Four publicly available cohorts of adults/children with viral infection, bacterial infection, | Influenza *vs* bacterial infection |

| | | | | | | |
|---|---|---|---|---|---|---|
| | | influenza, bacterial infection, non-infectious and healthy controls | | selected based on fold change and adjusted p value | non-infectious and healthy controls. One prospective cohort of adults presenting with suspected respiratory tract infection | |
| **IFI44L (9)** | NA | Children with viral or bacterial infection (6) | UK, USA and Spain | Elastic net followed by forward selection–partial least squares, using significantly differentially expressed transcripts | Children with bacterial or viral infection | Viral *vs* bacterial infection in febrile children |
| **IFIT3; RSAD2* (10)** | NA | Three cohorts of adults challenged with rhinovirus, influenza or RSV (11) | UK and USA | Sparse latent factor regression analysis on genome-wide data (11) followed by regularised logistic regression on the resulting 30-gene signature | Close contacts of students with acute upper respiratory viral infections | Pre-symptomatic viral infection *vs* healthy |
| **Li3 (12)** | Logistic regression (FS-PLS signature)§ | Adults with confirmed diagnosis of viral infection, bacterial infection or no infection | UK | Forward selection–partial least squares method (6) (13) applied to differentially expressed genes between definite bacterial and definite viral groups | One cohort of adults with definite viral, definite bacterial, probable viral, probable bacterial, indeterminate infection and non-infected/ other infection (e.g. fungal)<br>One cohort of adults with COVID-19 or confirmed bacteraemia | Viral (including COVID-19) *vs* bacterial |
| **Lopez7 (14)** | Sum of weighted gene expression values (bacterial *vs* viral classifier) | Children and adults with viral, bacterial, or non-infectious acute respiratory illness (15) | USA | Support vector machine analysis using genome-wide data | Children with acute viral or bacterial infections (16) | Viral *vs* bacterial respiratory infection |
| **Lydon15 (17)** | Logistic regression (Viral classifier)§ | Adolescents and adults with viral, bacterial, or non-infectious acute respiratory illness | USA | LASSO regression analysis using 87 selected target genes from previously derived signatures (15,18) | Patients with viral or bacterial co-infection or suspected bacterial infection | Viral *vs* bacterial respiratory infection |
| **MX1 (19)** | NA | NA | NA | Pre-selected due to biological plausibility | Adults challenged with the live yellow fever virus vaccine | Viral infection *vs* healthy |
| **Pennisi2 (20)** | Sum of downregulated genes subtracted from the sum of upregulated genes | Children with viral or bacterial infection (6) | UK, USA and Spain | Elastic net followed by forward selection–partial least squares, using significantly differentially expressed transcripts (6), then selection of an adequately expressed transcript for use in RT-LAMP | Children with bacterial or viral infection | Viral *vs* bacterial infection in children |
| **Rao8(21)** | Difference in geometric means between genes upregulated in bacterial infection & genes | Aggregated data from 32 publicly available cohorts containing subjects with viral infection, bacterial | Worldwide | Greedy backward search and abridged best subset selection performed on 100 genes with the highest scores in SAM analysis with LOSO analysis | Retrospective validation in aggregated data from 32 publicly available cohorts containing subjects with viral infection, bacterial infection and healthy | Viral *vs* bacterial infections |

| | | | | | | |
|---|---|---|---|---|---|---|
| | upregulated in viral infection (BoVI Score) | infection and healthy controls | | | controls and five individual cohorts containing subjects with viral infection and bacterial infection<br>Prospective validation in two cohorts of febrile adults & children with bacterial and viral infection | |
| **Ravichandran10 (22)** | Difference in geometric means between genes upregulated in bacterial infection and those downregulated in bacterial infection/upregulated in viral infection ($VB_{10}$) | Aggregated data from six publicly available cohorts containing subjects with viral infection, bacterial infection and healthy controls | UK, USA, Spain | Condition specific response networks computed from differentially expressed genes with network mining to identify top ranked perturbations from which top genes are selected based on a statistical threshold for differential gene expression across all discovery datasets | Aggregated data from 50 publicly available cohorts containing subjects with viral infection, bacterial infection and healthy controls<br><br>Bangalore – Viral Bacterial (BL-VB) cohort of adults with bacterial, viral and indeterminate infection and healthy controls | Viral *vs* bacterial infections |
| **RRM2(23)** | NA | Adults hospitalised with respiratory illness who tested positive or negative for COVID-19(24) | USA | Network analysis applied to differentially expressed genes and Maximal Clique Centrality (MCC) algorithm to identify top-ranked nodes | One cohort of adults with COVID-19 and healthy controls | COVID-19 *vs* healthy |
| **Sampson10 (25)** | SeptiCyte™ TRIAGE score (25) minus SeptiCyte™ VIRUS score (26) (Combined SeptiCyte score) | Eight cohorts of neonates, children, and adults with bacterial infections | UK, USA, Estonia and Australia | Regression analysis of transcript pairs using the 6000 most highly expressed genes from each dataset | Unselected consecutive patients presenting to the emergency department with febrile illness | Viral *vs* bacterial in febrile patients |
| **Sampson4 (26)** | Linear sum of upregulated and downregulated transcripts (Septicyte VIRUS) | Ten cohorts of children and adults with viral infections; two cohorts of adults challenged with influenza; and two cohorts of macaques challenged with Lassa virus or lymphocytic choriomeningitis virus | USA, Brazil, Finland and Australia | Regression analysis of transcript pairs using the 6000 most highly expressed genes from each dataset | Seven human cohorts and six non-human mammal cohorts infected or challenged with viruses across all seven of the Baltimore virus classification groups | Viral *vs* non-viral conditions |
| **Steinbrink19 (27)** | Logistic regression§ | Patients with candidaemia, viral infection, bacterial infection, or non-infectious SIRS and healthy controls | USA | Regularized multinomial logistic regression (LASSO) with nested leave one sample out cross-validation performed on differentially expressed genes identified by generalized linear hypothesis testing | Three cohorts: adults with candidaemia, viral infection, bacterial infection and healthy controls; children and adults with viral, bacterial or non-infectious acute respiratory illness, and healthy controls (15); children | Candidaemia *vs* bacterial *vs* viral infection *vs* SIRS *vs* healthy |

| | | | | | | |
|---|---|---|---|---|---|---|
| | | | | | with acute viral or bacterial infections (16) | |
| **Sweeney7 (28)** | Difference in geometric means between upregulated and downregulated genes, multiplied by ratio of counts of positive to negative genes (bacterial or viral metascore) | Eight cohorts of children and adults with viral and bacterial infections | USA, Australia, UK | Greedy forward search of 72 differentially expressed genes identified by multicohort analysis | 24 cohorts of children and adults with viral or bacterial infections, or healthy controls | Viral *vs* bacterial infection |
| **Trouillet-Assant6 (29)** | Median expression of 6 interferon-stimulated genes (Interferon score (30)) | NA | NA | Differential expression using 15 preselected interferon-stimulated genes | Febrile children with bacterial or viral infection | Viral *vs* bacterial infection in febrile children |
| **Tsalik33 (15)** | Logistic regression (Viral ARI classifier)§ | Children and adults with viral, bacterial, or non-infectious acute respiratory illness, and healthy controls | USA | LASSO regression analysis using the 40% of microarray probes with the largest variance after batch correction | Five cohorts of children or adults with viral, bacterial, or non-infectious respiratory illness, or viral or bacterial co-infection | Viral *vs* bacterial acute respiratory illness |
| **Xu2 (31)** | Logistic regression§ | Children and adults with acute febrile illness with confirmed bacterial infection, viral infection | China | Support vector machine learning to identify optimal combination of four candidate transcripts and binary logistic regression modelling | Children and adults with acute febrile illness with confirmed bacterial infection, viral infection, or non-infectious inflammatory disease | Viral *vs* bacterial infection |
| **Yu3 (8)** | Mean expression (non-RSV infections *vs* controls) | Children with acute respiratory illness and a positive result for a viral infection on a nasopharyngeal swab | USA | Modified supervised principal component analysis using all expressed transcripts | Children with RSV or rhinovirus infection | Viral *vs* healthy in children |
| **Zaas48 (18)** | Probit regression (Viral classifier)§ | Two cohorts of adults challenged with influenza A H3N2 or H1N1 | USA | Elastic net using 48 selected genes comprised of: 29 derived as a signature in a previous study (11) , seven shown to be downregulated in analysis of influenza challenge time course data (32) and 12 control genes | Adults presenting to the emergency department with fever and healthy controls | Viral vs bacterial acute respiratory illness |

Signatures are referred to by combining the first author's name of the corresponding publication as a prefix, with number of constituent genes as a suffix.

$Log_2$-transformed transcripts per million data used to calculate all signatures.

*Study by McClain et at sought to validate a 36-transcript signature for detection of respiratory viral infections. Model coefficients for the 36-transcript model are not provided; we therefore included the two best performing single transcripts from the study in the current analysis, since they demonstrated similar performance to the full model in the original publication.

[&]Where applicable, the name of the signature from the original publication is indicated in brackets.

§Logistic and probit regression models were calculated on the linear predictor scale using model coefficients from original publications.

[1]IFI27 is also identified as a marker of viral infection by Yu at al 2019 (8)

Acronyms: RSV= respiratory syncytial virus. PAM= prediction analysis of microarrays. LASSO=Least Absolute Shrinkage Selector Operator. RT-LAMP= Reverse Transcription Loop-mediated Isothermal Amplification. NA= not applicable. ATAC-seq= assay for transposase-accessible chromatin with sequencing. SAM= Significant Analysis of Microarray. LOSO= leave-one-study-out. SIRS= Systemic inflammatory response syndrome
